# Silent numerical failures in large language model–generated pharmacokinetic simulation code: a benchmark against target-controlled infusion validation criteria using the Marsh propofol model

**DOI:** 10.64898/2026.04.27.26351582

**Authors:** Masahito Omote

## Abstract

**Background:** Large language models (LLMs) are increasingly used by clinicians to generate executable code for pharmacokinetic (PK) simulation. Whether such code meets the accuracy standards of target-controlled infusion systems has not been systematically evaluated.

**Methods:** Five LLMs (ChatGPT, Claude, DeepSeek, Gemini, Grok) were prompted to generate Python code for the Marsh three-compartment propofol model under a standardized 120-minute bolus-plus-infusion regimen. Each LLM was tested in two phases: Phase 1, integrator free; Phase 2, fourth-order Runge–Kutta with 1-second step size mandated. Twenty runs per LLM per phase were collected (n = 200). Plasma concentrations were compared against a triple-validated reference using median prediction error (MDPE), median absolute prediction error (MDAPE), and Wobble. Runs were classified as Class A (MDAPE < 1 %), B (1–30 %), C (≥ 30 %), or D (failed).

**Results:** All 200 scripts were invokable and created a CSV file; 199/200 (99.5 %, 95 % CI 97.1–99.9 %) produced a valid time–concentration series. The remaining script (Gemini Phase 2 run 18) aborted during row formatting with `ValueError` and left a header-only CSV. Median MDAPE per LLM × phase ranged 0.0043–0.020 %, with 195/200 runs (97.5 %, 95 % CI 94.3–98.9 %) achieving Class A. Five runs (2.5 %, 95 % CI 1.1–5.7 %) were non-excellent or structurally defective: three were Class C due to time-scale/unit-handling errors (one DeepSeek run with a 6-second effective Euler step from a minute-as-second declaration, two Grok runs with min⁻¹ rate constants applied per second), one was Class D (the empty-CSV failure above), and one was Class B but reflected a duplicated-bolus implementation error rather than a benign numerical deviation. Kruskal–Wallis testing showed significant inter-LLM heterogeneity across all metrics and phases (all omnibus p < 0.01). Strict compliance with Phase 2 directives was 98 % (98/100 runs; 95 % CI 93.1–99.5 %); lenient compliance accepting RK4-adaptive implementations as a superset was 100 % (100/100 runs; 95 % CI 96.3–100 %). Yet all three numerically divergent Phase 2 runs occurred under nominally compliant RK4/dt = 1 s configurations; the fourth non-Class-A Phase 2 outcome was a formatting failure that produced no usable trajectory.

**Conclusion:** LLMs generate numerically accurate Marsh-model code in most runs but silently diverge in a clinically non-negligible minority. The Marsh model — the simplest fixed-parameter three-compartment propofol model — functioned here as a positive control: even so, three distinct classes of structural bug (unit/time-scale mismatch, duplicated-bolus event handling, malformed f-string formatting) slipped past apparent execution success. Two additional Phase 2 runs used an RK4-adaptive variant rather than classical RK4 and are therefore better interpreted as strict prompt non-compliance than as numerical failure. Prompt-level method specification substantially reduced algorithm-selection errors but did not eliminate unit or structural bugs. LLM-generated pharmacokinetic code requires reference-based validation before any safety-relevant use.

## 1. Introduction

Large language models (LLMs) have rapidly entered clinical and biomedical workflows. Beyond literature review and clinical decision support, physicians and researchers increasingly prompt LLMs to generate executable code for quantitative biomedical tasks, and much of that code is then run directly, without independent verification, to inform analysis or (in the limit) dosing. The safety question this raises is distinct from the question traditionally asked of LLMs in medicine — whether their natural-language output is factually correct. Code does not need to be factually correct to execute; it needs to execute *and* to produce numerically faithful output. An LLM-generated script can read plausibly, import successfully, run to completion, and write a well-formed output file, and yet encode a silent structural bug that renders its numerical output clinically unusable. The end user, seeing a file on disk, may have no indication that anything went wrong.

In anesthesiology, this combination of “runs cleanly, output is wrong” is particularly consequential. Pharmacokinetic (PK) modeling underlies the safe administration of intravenous anesthetics, particularly in target-controlled infusion (TCI) systems, which must predict plasma and effect-site concentrations to schedule bolus doses and maintain target concentrations. Existing validated clinical tools (e.g., Tivatrainer) rely on carefully implemented multi-compartment PK models; any ad-hoc simulator — including an LLM-generated one — is downstream of the same numerical-methods requirements, but without the human review path. A clinician who asks an LLM to “write a Python script that simulates propofol using the Marsh model” is, in effect, commissioning safety-relevant pharmacokinetic software in a single turn, with no specification review, no code review, and no reference testing.

The methodological challenge for evaluating such code is that apparent success — syntactically valid code that executes and writes a file — is not itself evidence of numerical fidelity. Neither is visual inspection of a plasma concentration curve: sub-percent drift is invisible to the eye, and a wrong-but-plausible curve can be clinically unusable even when it looks reasonable. A quantitative benchmark is needed. In clinical TCI validation, the traditional metrics are the median prediction error (MDPE), median absolute prediction error (MDAPE), and Wobble of Varvel et al. [11]; these are the criteria against which commercial TCI systems have been validated for decades, and they are directly applicable to the outputs of LLM-generated PK code.

Recent studies have begun to explore the application of LLMs to pharmacometrics workflows, though with a focus on domain-specific modeling platforms rather than standalone simulation code. Shin et al. evaluated ChatGPT and Gemini on NONMEM coding tasks and found that while the models could generate initial coding templates, the output contained structural and syntax errors requiring expert revision [1]. Cloesmeijer et al. investigated ChatGPT’s ability to generate R scripts for a one-compartment population PK model with allometric scaling and develop a Shiny-based interface for dosing simulations, reporting that occasional coding errors and task misinterpretation necessitated careful human oversight [2]. More recently, Cha et al. compared four LLMs (GPT-4o, Claude 3.5 Sonnet, Gemini 1.5 Pro, and Llama 3.2) on NONMEM output interpretation and simulation tasks; ChatGPT successfully simulated a three-compartment meropenem model with pharmacokinetic parameter differences of less than 1 % compared to NONMEM, but encountered difficulty with a more complex two-compartment nivolumab model incorporating multiple covariates, where incomplete covariate implementation led to larger discrepancies [3]. Sánchez Herrero and Calvet compared ChatGPT 3.5, Microsoft Copilot v4.0, and Google Bard/Gemini on the construction of a two-compartment population PK model in R, and reported small parameter-level differences relative to a reference model (AFE 0.99, AAFE 1.14, MPE −1.85), with ChatGPT and Copilot outperforming Gemini on qualitative coding tasks [4]. In a parallel line of work, Holt et al. demonstrated that LLMs can contribute to the data-driven discovery of dynamical systems in pharmacology, using LLMs as scientific-reasoning agents rather than as code-writers for a pre-specified PK model [5]. Most recently, Zheng et al. evaluated seven OpenAI-family LLMs (including the OpenAI reasoning model `o1` and GPT-4.1) on 13 NONMEM pharmacometric coding tasks with a rubric-based scoring scheme, showing that `o1` and GPT-4.1 achieved the best overall performance, that increases in model complexity (indirect-response and absorption-lag models) degraded performance in the baseline condition, and that an “optimized prompt” substantially improved accuracy across all models [6]. Collectively, these studies establish that LLM-based assistance with pharmacometric modeling is feasible for certain tasks and that prompt-level guidance can move the distribution of generated code toward better outputs; however, none of these prior works have systematically evaluated whether LLM-generated, standalone, directly executable simulation code for a named clinical PK model meets the quantitative validation criteria used for target-controlled infusion (TCI) systems, nor has any prior study characterized the distribution of such code across repeated independent draws from multiple contemporary LLMs spanning several providers.

A critical gap remains: no study has evaluated LLM-generated PK simulation code using the quantitative criteria routinely applied in pharmacokinetic model validation. Existing evaluations have relied primarily on expert code review, visual comparison of concentration–time profiles, or non-compartmental analysis of simulated outputs. Clinically established performance metrics such as prediction error (PE), median prediction error (MDPE), and median absolute prediction error (MDAPE), which are the standard for assessing the accuracy of TCI systems and PK model implementations, have not been applied to LLM-generated code.

Pharmacokinetic models for propofol offer an ideal benchmark for this evaluation. The three-compartment mammillary model has well-established parameter sets of graded complexity: the Marsh model [7] uses fixed PK parameters, the Schnider model [8] introduces covariate-dependent parameters based on patient demographics, and the Eleveld model [9] incorporates a substantially more complex mathematical structure with additional covariates and allometric scaling. Validated reference implementations and the established clinical tool Tivatrainer provide a rigorous ground truth against which LLM-generated code can be compared.

In this study we evaluate the runtime numerical output of LLM-generated pharmacokinetic simulation code against clinically established TCI validation criteria, using the Marsh three-compartment propofol model as a deliberately simple, fixed-parameter, widely-implemented positive control. The choice of Marsh is motivated by the fact that it is the simplest of the propofol PK models routinely used in clinical TCI (fixed, covariate-free parameters; well-documented reference implementations; no allometric scaling or effect-site component in its baseline form); if contemporary LLMs were going to fail on any compartmental propofol task, this is the task we would expect them to fail least often. A high failure rate on Marsh would therefore be a prima-facie signal that the current generation of LLMs is not ready to produce safety-relevant PK code unsupervised. A low failure rate does not establish safety, but the residual failures — if any — provide a high-value taxonomy of the failure modes that clinicians and reviewers of LLM-generated medical code should expect to encounter.

We therefore ask three concrete questions. (i) In the aggregate, how numerically accurate is LLM-generated Marsh-model code when evaluated against a validated reference using the TCI metrics? (ii) Does the fraction of clinically unacceptable output change when the prompt explicitly specifies the numerical integration method and step size (a zero-cost intervention available to any user)? (iii) When a generated script does fail — whether by producing a clinically unacceptable trajectory, by producing no usable output at all, or by producing a nominally Varvel-acceptable trajectory via a structural implementation bug — what are the failure modes, and are they predictable and detectable from inspection of the generated source? To our knowledge this is the first systematic evaluation of LLM-generated, directly executable pharmacokinetic simulation code for the Marsh propofol model using clinically established TCI validation metrics (MDPE, MDAPE, Wobble) applied to the runtime numerical output of the generated code, with repeated independent trials per LLM across five contemporary LLMs from five different providers.

## 2. Methods

### 2.1. Study design

This study was an observational, simulation-based evaluation of pharmacokinetic code generation by contemporary large language models. No patient data, human subjects, or animal experiments were involved. Because the study consisted entirely of in silico simulations executed against a synthetic reference dataset, and involved no human participants, animal subjects, or identifiable patient data, institutional review board approval was not required.

We evaluated five contemporary LLMs available to end users through publicly accessible chat interfaces: **ChatGPT** (GPT-5.3 Instant, with auto-route disabled; OpenAI, Inc., San Francisco, CA, USA), **Claude** (Claude Sonnet 4.6; Anthropic PBC, San Francisco, CA, USA), **DeepSeek** (DeepSeek-V3.2 in Instant mode; DeepSeek, Hangzhou, China), **Gemini** (Gemini 3, displayed as “Fast” in the web UI; Google LLC, Mountain View, CA, USA), and **Grok** (Grok 4.1 Fast; xAI Corp., Palo Alto, CA, USA). All five LLMs were accessed through the vendor’s public chat interface (no API access) using the default non-reasoning response mode; slower “Thinking” / “Reasoning” variants that each vendor also makes available were deliberately not used, to reflect the most common end-user interaction pattern (see §4.5). The exact model strings, access dates, access tier (free versus paid), and account configurations for each LLM are summarized in Supplementary Table S1 and in the run-level metadata provided in the open data release (see Data Availability).

The evaluation focused on the Marsh three-compartment pharmacokinetic model for propofol [7]. This model was selected as a deliberate *positive control* for the LLM code-generation task, because (i) its parameters are fixed (independent of patient covariates), which eliminates covariate-implementation as a confounding source of error; (ii) it is one of the most widely implemented PK models in target-controlled infusion devices and research simulators, so its structural equations are highly represented in the training-data distribution of any current LLM; and (iii) validated reference implementations are available for ground-truth comparison. We expect the Marsh task to be easier for current LLMs than the covariate-dependent Schnider model or the allometrically-scaled Eleveld model; any failure rate observed here is therefore a lower bound on the failure rate that should be expected for more complex propofol models and, by extension, for other compartmental PK tasks. Evaluation of Schnider and Eleveld is planned for subsequent work.

### 2.2. Standardized prompt and two-phase protocol

A single standardized natural-language prompt was developed and used identically across all LLMs and all runs. The prompt specified: (1) the task (simulate the plasma concentration of propofol using the Marsh three-compartment pharmacokinetic model); (2) the programming language (Python); (3) an explicit prohibition on external ODE solver libraries (`scipy.integrate.odeint`, `scipy.integrate.solve_ivp`, and similar), requiring each LLM to implement the numerical integrator from scratch; (4) the patient (70 kg adult); (5) the Marsh model parameters (V_c_ = 228 mL·kg⁻¹; k_10_ = 0.119, k_12_ = 0.112, k_13_ = 0.0419, k_21_ = 0.055, k_31_ = 0.0033 min⁻¹), with the 1991 primary source cited [7]; (6) the dosing schedule (bolus 140 mg into the central compartment at t = 0 min; continuous infusion of 700 mg·h⁻¹ from 0–10 min, 560 mg·h⁻¹ from 10–20 min, 420 mg·h⁻¹ from 20–60 min, and 0 mg·h⁻¹ from 60–120 min); (7) a simulation horizon of 0–120 min; and (8) the required CSV output format (two columns `time_sec` and `Cp` in µg·mL⁻¹, at 1-second intervals).

Crucially, the prompt did **not** specify the numerical integration algorithm, internal time step, or any other implementation detail beyond the output format and the prohibition on external solvers. The full prompt text (both the Phase 1 and Phase 2 versions) is provided verbatim in Supplementary Material S2.

The prompt was deliberately designed to probe two distinct implementation burdens that commonly cause silent errors in scientific computing. First, the micro-rate constants were supplied in min⁻¹ (the canonical clinical unit used in the original 1991 Marsh publication) while the required output sampling was at 1-second intervals, so that a minute-to-second conversion was an obligatory implementation step that every LLM had to perform correctly in its own integration loop. Second, the central volume was stated as V_c_ = 228 mL·kg⁻¹ rather than as a pre-computed V_1_ = 15.96 L, obliging each LLM to perform the body-weight multiplication and the milliliter-to-liter conversion independently. These prompt-level choices were made so that unit-handling — a well-known source of error in hand-written pharmacokinetic code, and a plausible source of silent error in LLM-generated code — was tested as part of the evaluation rather than abstracted away by the specification. The minute/second mismatch that produced two of the three numerically divergent runs in the present dataset (§3.6) is therefore the detection, not the discovery, of a failure mode deliberately included in the evaluation’s scope.

The protocol consisted of two phases:

- **Phase 1 (integrator free).** The standard prompt was used without modification. LLMs were free to choose any numerical integration method and internal step size.
- **Phase 2 (RK4 constrained).** A single additional requirement was inserted into the prompt, mandating the classical fourth-order Runge–Kutta (RK4) method with a fixed time step of dt = 1 second.

For each LLM and each phase, 20 independent runs were collected (total n = 200 runs). Each run corresponded to a single fresh conversation: to minimize carry-over and in-context learning effects, every run was initiated in a new chat session, and no clarifying or follow-up messages were exchanged beyond the prompt. The code returned by the LLM was executed without modification in a standard Python 3 environment; only obvious non-code artifacts such as surrounding prose or markdown fences were stripped prior to execution. Any execution error (syntax error, runtime exception, non-terminating output, or empty output file) was recorded as such in the outcome analysis rather than re-prompted or fixed.

Each generated code file, its SHA-256 hash, its stdout/stderr, and the produced CSV were archived at the time of collection to ensure byte-level reproducibility. All SHA-256 hashes are recorded in the per-run `metadata.json` files included in the open dataset release (see Data Availability). LLM-generated code was executed without modification in a standard Python 3 environment (Python 3.11 on Windows 11 Pro 24H2).

### 2.3. Reference implementation and ground truth

We adopt a deliberately minimal author-implemented reference simulator as Ground Truth, rather than a mature domain-specific PK package such as `rxode2` or `nlmixr`. This choice reflects three considerations. First, for the Marsh model — a fixed-parameter three-compartment mammillary system with no covariate dependence and no absorption phase — the numerical task reduces to tightly-bounded linear ODE integration, for which a well-validated hand-written RK4 implementation at dt = 0.01 s is numerically indistinguishable from any general-purpose solver. Second, the comparison being made here is not between two modeling platforms but between LLM-generated code and a reference trajectory; introducing an additional layer of platform-specific abstraction (e.g., rxode2’s event-table formalism) would obscure rather than clarify what is being measured. Third, the reference simulator is itself released as part of the open-source repository supporting this paper, so that the Ground Truth is transparently inspectable and independently auditable by readers.

A reference simulator for the Marsh model was implemented independently by the author in plain Python. The central (V_1_ = 228 mL·kg⁻¹ × 70 kg = 15.96 L), peripheral, and deep peripheral compartments were modeled as a three-compartment mammillary system, with amounts A_1_, A_2_, A_3_ (mg) in each compartment governed by the ordinary differential equation (ODE) system

- dA_1_/dt = I(t) − (k_10_ + k_12_ + k_13_) A_1_ + k_21_ A_2_ + k_31_ A_3_
- dA_2_/dt = k_12_ A_1_ − k_21_ A_2_
- dA_3_/dt = k_13_ A_1_ − k_31_ A_3_

where I(t) is the infusion rate (mg·s⁻¹) at time t and all micro-rate constants have been converted from min⁻¹ to s⁻¹. A 140 mg bolus was applied as an instantaneous initial condition A_1_(0) = 140 mg, A_2_(0) = A_3_(0) = 0. The ODE system was integrated with a hand-written classical fourth-order Runge–Kutta (RK4) scheme at a fixed internal step size of 0.01 s over 0–7200 s, with the simulated plasma concentration C_p_(t) = A_1_(t) / V_1_ sampled and recorded at 1-second intervals to yield 7201 reference points directly comparable to the LLM-generated CSV outputs.

The reference implementation was triple-validated. **First**, the hand-written RK4 solution was cross-checked against `scipy.integrate.odeint` solving the same three-compartment system with `rtol` `=` `atol` `=` `1` `×` `10⁻¹²`, and agreement was within a maximum relative deviation of 4.4 × 10⁻⁴ % across the full 0–7,200 s horizon. **Second**, output from the widely used clinical simulator Tivatrainer (Pro edition, monthly subscription; version 2.2.0, Build 287; Gutta BV, Aerdenhout, The Netherlands; available at https://www.tivatrainer.com, accessed April 2026), run on the same host operating system [10], was compared against the reference under an identical dosing regimen. The Tivatrainer user interface does not permit a bolus and an infusion to be registered at exactly the same time point; as a consequence, the Tivatrainer infusion schedule is internally shifted by 1 s relative to the bolus (bolus at t = 0, infusion onset at t = 1 s, and all subsequent infusion transitions similarly shifted to t = 601 s, 1201 s, and 3601 s). A dedicated “Tivatrainer-matched” mode of the reference simulator (`--tivatrainer`) reproduces this offset exactly; under that mode, the reference and Tivatrainer outputs agreed to within 0.06 % median absolute prediction error across all time points from t = 1 s to t = 7200 s. **Third**, all Marsh parameters were verified against the original 1991 publication [7], and the central-volume unit conversion (228 mL·kg⁻¹ × 70 kg = 15.96 L) was independently re-derived.

For all subsequent comparisons against LLM-generated runs, the “standard” mode of the reference simulator (infusion onset at t = 0 s, as explicitly specified in the prompt and as all LLMs implemented it) was used as the ground truth.

### 2.4. Accuracy metrics

For each LLM-generated run, the predicted plasma concentration C_pred_(t) was compared against the reference plasma concentration C_ref_(t) at every second from t = 0 to t = 7200 s (n = 7201 time points). The prediction error at time t was defined in the standard pharmacokinetic form:

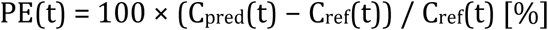

From the per-run PE series we derived three summary metrics widely used in TCI system validation [11]:

- **MDPE** (median prediction error): median of PE(t) across all time points — a measure of systematic bias.
- **MDAPE** (median absolute prediction error): median of |PE(t)| — a measure of inaccuracy.
- **Wobble**: median absolute deviation of PE(t) from the run’s MDPE — a measure of intra-run variability.

The per-run metrics were then aggregated across the 20 runs per LLM × phase cell. Samples at C_ref_(t) = 0 were excluded from PE calculation; no such samples occurred in practice.

### 2.5. Classification by clinical pharmacokinetic criteria

Each run was classified into one of four ordinal categories:

- **Class A (excellent):** MDAPE < 1 %, indicating numerical reproduction of the reference.
- **Class B (clinically acceptable):** 1 % ≤ MDAPE < 30 %, satisfying the Varvel criterion traditionally used to validate TCI systems [11].
- **Class C (clinically unacceptable):** MDAPE ≥ 30 %, exceeding the conventional clinical acceptance threshold.
- **Class D (execution failure):** code did not execute successfully or did not produce a valid output CSV.

### 2.6. Step size and algorithm analysis

To characterize the choices LLMs made when given free discretion (Phase 1) and the extent to which they complied with explicit directives (Phase 2), the Python code returned for each of the 200 runs was parsed programmatically. Assignments to candidate step-size variables (dt, dt_sec, dt_min, h, step_size, and case variants) were extracted using Python’s ast module, including default values in function definitions. Simple arithmetic expressions (e.g., 1.0 / 60.0) were evaluated symbolically; cross-variable references were resolved through an incrementally built variable-context map. Units were inferred from a combination of variable-name affixes (_sec, _min), in-line comments, and structural cues in the assignment expression (/ 60, * 60), and all step sizes were normalized to seconds for comparison. The integration algorithm actually implemented (Euler, RK4, or adaptive variants) was determined by manual inspection of the integration loop. Full step-size extraction results including the source expression, inferred unit, and normalized value are provided for every run in the supplementary data.

For Phase 2 compliance analysis, we report two complementary rates: **strict compliance** counts only runs that implemented the classical fourth-order Runge–Kutta scheme exactly as specified in the prompt; **lenient compliance** additionally counts runs that implemented an adaptive or otherwise augmented RK4 variant (e.g., RK4 with adaptive step-size correction) as a strict superset of the requested algorithm. The distinction is relevant because the two DeepSeek Phase 2 runs (`p2_run17` and `p2_run19`) self-described their implementation as “RK4 with adaptive step-size correction” rather than classical RK4; both produced numerically excellent output (MDAPE < 0.01 %) and are therefore scored as compliant under the lenient definition but not under the strict definition.

### 2.7. Statistical analysis

The pre-specified primary outcome of this study is the pooled proportion of runs across all 5 LLMs and both phases that produced clinically unacceptable numerical output or execution failure (Class C + D), reported with a two-sided 95 % Wilson score confidence interval. Secondary outcomes are (i) the phase-pooled proportion of runs exhibiting any structural defect (including Class B runs with identifiable implementation bugs), (ii) the proportion of Phase 2 runs complying with the RK4 / dt = 1 s directive, and (iii) the per-LLM × per-phase distributions of MDPE, MDAPE, and Wobble. Per-LLM comparisons of failure rate and of metric distribution are treated throughout as exploratory: at n = 20 runs per LLM per phase, the study is not powered to establish provider-level ordering of failure risk, and all between-LLM differences reported are interpreted as hypothesis-generating for future work with larger per-LLM sample sizes.

Because the metrics distributions were heavy-tailed and included occasional divergent outliers, all group comparisons were performed with non-parametric tests. MDPE, MDAPE, and Wobble distributions were compared across the five LLMs within each phase using the Kruskal–Wallis H test. When the omnibus Kruskal–Wallis test indicated heterogeneity, pairwise post-hoc comparisons between LLMs were performed using Dunn’s test with Bonferroni correction for the 10 pairwise comparisons per phase × metric cell, implemented in the scikit-posthocs library (p_adjust=“bonferroni”). The p-values returned by this procedure are already Bonferroni-adjusted, so we judge significance against the conventional thresholds α = 0.05 (marked *) and α = 0.01 (marked **), not against further-corrected values. Within-LLM comparisons of Phase 1 versus Phase 2 were performed with the Mann–Whitney U test (two-sided), treating the two phases as independent samples since the runs were independently generated; because each metric involved 5 such within-LLM comparisons (one per LLM), we multiplied the raw two-sided p-values by 5 to obtain Bonferroni-adjusted p-values, and again judged significance against α = 0.05 (*) and α = 0.01 (**). In both cases the reported p-values are adjusted p-values; “n.s.” denotes an adjusted p-value that did not reach α = 0.05.

Descriptive statistics are reported as median [interquartile range, IQR] throughout. Proportions are reported with two-sided 95 % Wilson score confidence intervals. Because each run was initiated in a fresh, independent chat session with no shared conversational state, runs from the same LLM were considered statistically independent. Quantitative accuracy comparisons (MDPE, MDAPE, Wobble; Figure 1, Figure 4, and Supplementary Figure S2) are conditional on the run having produced a valid time–concentration CSV, so a single execution failure (Gemini `p2_run18`) is excluded from these analyses but is retained in the ordinal reliability classification (Class A/B/C/D; Figure 2 and §3.5) as a Class D outcome. This separation is explicit by design: the metric-based analyses characterize numerical fidelity when the generated code does produce output, while the Class A/B/C/D summary characterizes end-to-end reliability including the possibility of producing no usable output at all. All analyses were performed in Python 3 using scipy.stats (v ≥ 1.10) and scikit-posthocs (v ≥ 0.7); plots were produced with matplotlib.

**Figure 1.**
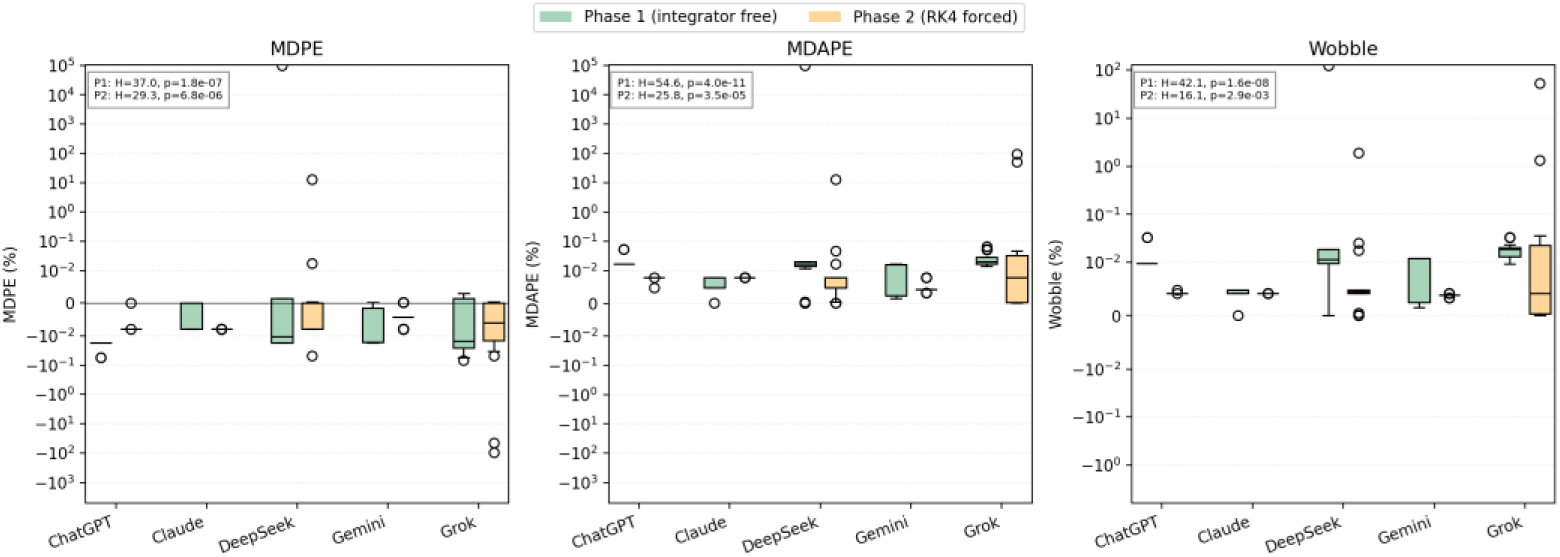
Performance metrics of LLM-generated Marsh-model code across five LLMs and two prompt phases. Box-and-jitter plots of the three pharmacokinetic accuracy metrics — median prediction error (MDPE, top row), median absolute prediction error (MDAPE, middle row), and Wobble (bottom row) — stratified by LLM (x-axis) and phase (columns: Phase 1, integrator free; Phase 2, fourth-order Runge–Kutta with dt = 1 s mandated). Each dot represents one independent code-generation run (n = 20 per LLM per phase, except Gemini Phase 2 where n = 19 after exclusion of one execution failure). Boxes show median and interquartile range; whiskers extend to the 1.5 × IQR limits. The y-axis uses a symmetric-logarithmic scale (linear threshold ± 0.01 %) to visualize both the tight central distributions (sub-percent) and the extreme divergent runs simultaneously. Inset boxes report the Kruskal–Wallis H statistic and p-value for the test of LLM-level heterogeneity within each panel; all six tests were significant at p < 0.01 (**).

**Figure 2.**
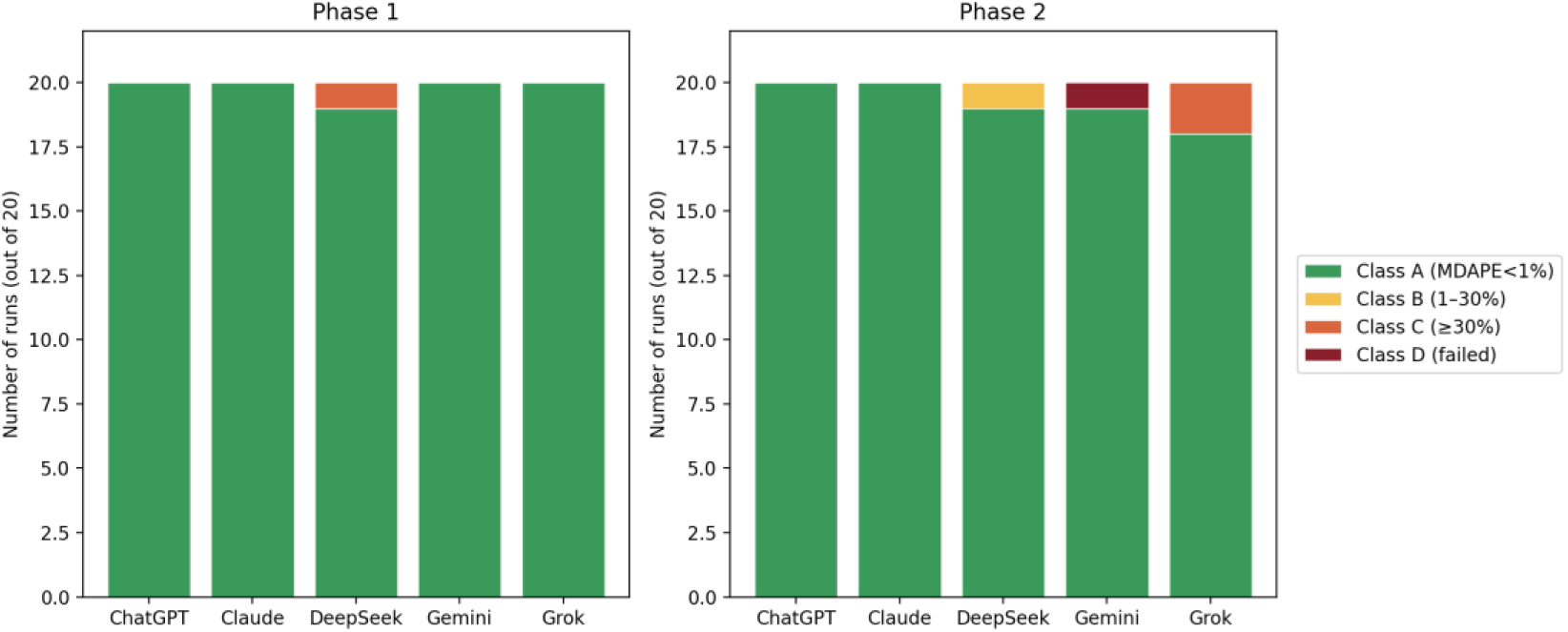
Classification of LLM-generated runs by clinical pharmacokinetic criteria. Stacked bar plot showing the classification of each of the 20 runs per LLM per phase (Phase 1, left panel; Phase 2, right panel) into four ordinal categories based on median absolute prediction error (MDAPE) and execution status: Class A (green, MDAPE < 1 %); Class B (yellow, 1 % ≤ MDAPE < 30 %, Varvel-acceptable [11]); Class C (orange, MDAPE ≥ 30 %); Class D (dark red, execution failure). Numbers on each colored segment indicate the count of runs in that class.

**Figure 3.**
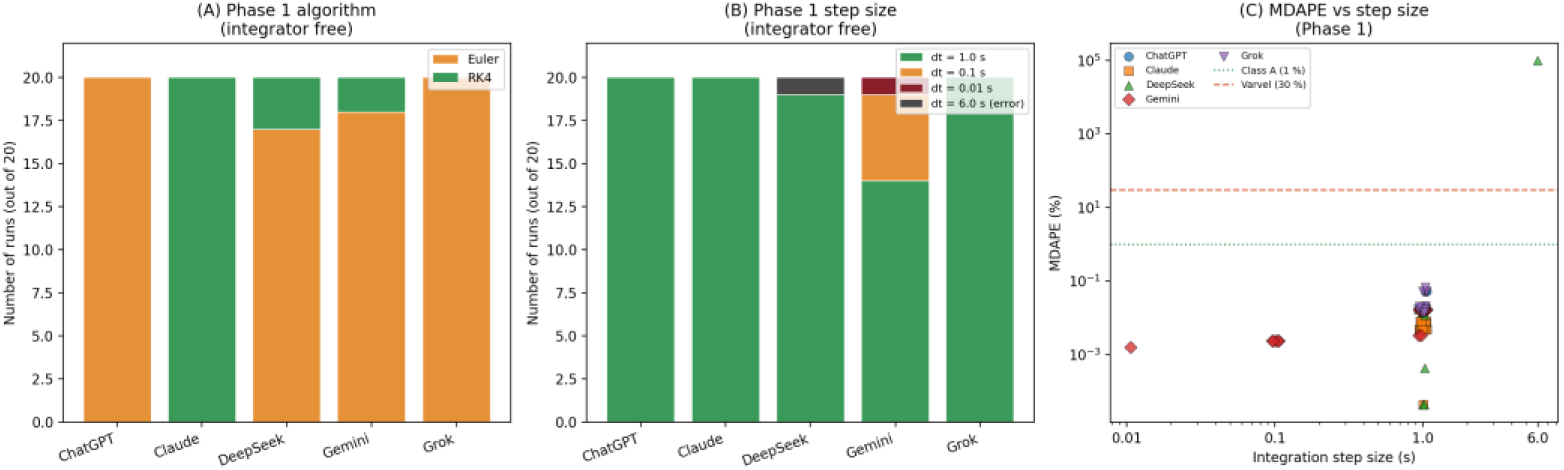
Integration algorithm and step size chosen by LLMs, and their effect on accuracy. (A) Phase 1 algorithm distribution (integrator free). Stacked bar plot showing the distribution of integration algorithm across the 20 Phase 1 runs for each LLM: Euler (orange) versus classical fourth-order Runge–Kutta (RK4, green). **(B) Phase 1 step-size distribution.** Stacked bar plot of the internal integration step size after unit normalization to seconds: dt = 1.0 s (green), dt = 0.1 s (orange), dt = 0.01 s (dark red), and dt = 6.0 s (dark gray, arising from a unit misinterpretation in DeepSeek `p1_run05`). Gemini was the only LLM to self-select sub-second step sizes. **(C) MDAPE per run versus step size (Phase 1 only).** Jittered scatter plot of per-run MDAPE against the inferred internal step size. Horizontal reference lines indicate the Class A threshold (MDAPE = 1 %, green dotted) and the Varvel clinical-acceptance criterion (MDAPE = 30 %,red dashed). The single divergent Phase 1 run (DeepSeek `p1_run05`, MDAPE ≈ 10⁵ %) sits far above the Varvel line at dt = 6 s, while the six Gemini sub-second runs produced the lowest per-run MDAPE values observed in the dataset. In Phase 2, all 100 runs used dt = 1 s and either classical RK4 (98/100) or an RK4-adaptive variant (2/100); strict classical-RK4 compliance was 98 %, and lenient RK4-family compliance was 100 %. The corresponding distribution is flat and not shown.

**Figure 4.**
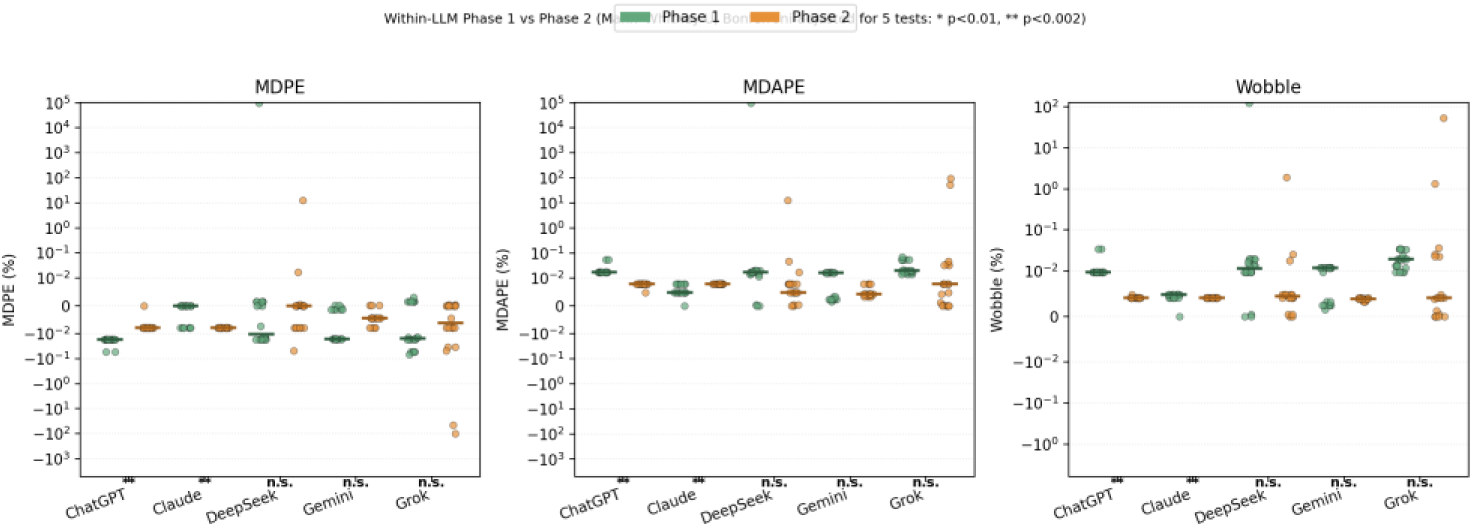
Within-LLM comparison of accuracy metrics between Phase 1 (integrator free) and Phase 2 (RK4 forced, dt = 1 s mandated). Three-panel box-and-jitter plot comparing Phase 1 (green) and Phase 2 (orange) distributions of MDPE (left), MDAPE (middle), and Wobble (right) for each LLM. Each dot is one run (n = 20 per cell, Gemini Phase 2 n = 19). Short horizontal bars indicate the within-group medians. Y-axis uses symmetric-logarithmic scale. Significance markers below each LLM indicate the Bonferroni-adjusted Mann–Whitney U two-sided p-value (raw p × 5) for the within-LLM Phase 1 vs Phase 2 comparison, judged against α = 0.05 for *, 0.01 for **, with “n.s.” denoting p_adj_ ≥ 0.05.

### 2.8. Use of AI tools

In addition to the LLMs being themselves the subject of evaluation, the author used a large language model (Claude, Anthropic PBC) to assist with code review, data analysis workflow, and manuscript drafting. All scientific claims, numerical results, and the final text were verified and approved by the author, who takes full responsibility for the integrity of the work.

## 3. Results

### 3.1. Overview of the dataset

Across five contemporary LLMs and two phases, a total of 200 runs were generated (20 runs per LLM × phase cell). All 200 scripts were invokable, and all 200 created a CSV output file on disk. In one case, however — Gemini Phase 2 run 18 — the CSV contained only the header row and no data, because the script raised a `ValueError` during row formatting and aborted the write immediately after the header line was emitted (see §3.6 for details). We therefore report 199 runs (99.5 %, 95 % CI 97.1–99.9 %) as yielding a valid plasma-concentration time series over the full 0–7200 s horizon, and 1 run (0.5 %) as an execution failure for the purposes of accuracy evaluation.

All 199 runs that produced a valid CSV contained the required two-column structure (time_sec, Cp) and 7201 one-second samples. Because the prediction error was evaluated at every sample, each of these runs contributed 7201 PE values, and no run required exclusion due to zero reference concentrations.

### 3.2. Overall accuracy across LLMs and phases

The distribution of the three accuracy metrics — MDPE, MDAPE, and Wobble — is shown in Figure 1, stratified by LLM and phase. In both phases, Kruskal–Wallis testing indicated strong heterogeneity across LLMs for all three metrics (Phase 1: H = 37.0, 54.6, and 42.1 with p = 1.8 × 10⁻⁷, 4.0 × 10⁻¹¹, and 1.6 × 10⁻⁸ for MDPE, MDAPE, and Wobble, respectively; Phase 2: H = 29.3, 25.8, and 16.1 with p = 6.8 × 10⁻⁶, 3.5 × 10⁻⁵, and 2.9 × 10⁻³).

Median MDAPE (the primary accuracy measure) in Phase 1 ranged from 0.0048 % for Claude to 0.0202 % for Grok, with all per-LLM medians below 0.025 %. In Phase 2 under RK4 with a 1 s step size, median MDAPE ranged from 0.0043 % for Gemini to 0.0079 % for ChatGPT, Claude, and Grok. Thus, for the majority of runs, every LLM produced code whose plasma-concentration profile agreed with the reference to within a small fraction of one percent — well below any clinically meaningful threshold.

However, the Kruskal–Wallis result is driven only partly by these central tendencies; it is substantially amplified by a small number of divergent runs. Per-run MDAPE values reached 99,762 % (DeepSeek Phase 1 run 05), 12.9 % (DeepSeek Phase 2 run 14), and 97.9 % (Grok Phase 2 run 13), compared with per-run maxima below 0.07 % for ChatGPT, Claude and Gemini in both phases (Figure 1, outliers visible on the symmetric-log axis). For these catastrophically divergent runs the absolute MDPE and MDAPE values coincide because every per-time-point error has the same sign. The nature and implementation-level origin of these divergent runs is examined below.

### 3.3. Pairwise LLM comparisons

Pairwise Dunn’s tests with Bonferroni correction between the five LLMs within each phase are summarized as p-value matrices in Supplementary Figure S2. The p-values reported below are already Bonferroni-adjusted; we judge significance against the conventional thresholds α = 0.05 (*) and α = 0.01 (**). The clearest separations were:

In Phase 1, Claude was significantly separated from ChatGPT on MDAPE (p = 4.7 × 10⁻⁷), from Grok on MDAPE (p = 6.2 × 10⁻⁸) and Wobble (p = 4.7 × 10⁻⁹), and from DeepSeek on MDAPE (p = 9.3 × 10⁻⁴) and Wobble (p = 5.1 × 10⁻⁴). Gemini was significantly separated from ChatGPT on MDAPE (p = 1.0 × 10⁻⁴) and from Grok on both MDAPE (p = 1.9 × 10⁻⁵) and Wobble (p = 1.0 × 10⁻³), reflecting the influence of its sub-second step-size choices (§3.6). In Phase 2, Gemini was separated from ChatGPT (MDAPE p = 8.1 × 10⁻⁵) and from Claude (MDAPE p = 4.6 × 10⁻⁴), but these Phase 2 differences no longer reached the Bonferroni-adjusted threshold on Wobble (ChatGPT vs Gemini p = 9.4 × 10⁻², Claude vs Gemini p = 2.7 × 10⁻²), indicating that the Phase 2 inter-LLM differences are MDAPE-dominated rather than Wobble- dominated. ChatGPT and Claude did not show a statistically significant difference from one another on any metric in Phase 2 (MDAPE p = 1.00 after Bonferroni correction); we cannot conclude from this that the two distributions are equivalent, only that our sample was insufficient to detect a difference if one exists. Grok’s Phase 2 distribution was not significantly separated from other LLMs after Bonferroni correction, despite raw Mann–Whitney p-values below 0.05 in several comparisons, consistent with the two divergent runs (§3.6) inflating its variance without shifting its central tendency.

The full pairwise matrix, including all Dunn–Bonferroni adjusted p-values, is provided in Supplementary Figure S2.

### 3.4. Effect of forcing RK4 and a fixed step size

Phase 2 added a single sentence to the prompt mandating RK4 integration with dt = 1 s. Within-LLM comparisons of Phase 1 versus Phase 2 (Figure 4) revealed heterogeneous effects. The p-values reported below are Bonferroni-adjusted Mann–Whitney U two-sided p-values (raw p × 5 to correct for the 5 within-LLM tests per metric), judged against α = 0.05 (*) and α = 0.01 (**).

**ChatGPT** showed the largest shift, with all three metrics significantly changing from Phase 1 to Phase 2 (all Bonferroni-adjusted p = 1.2 × 10⁻⁸). Median MDAPE decreased from 0.017 % to 0.0079 %, reflecting the replacement of Euler integration (used in 20/20 Phase 1 runs) by RK4 (20/20 Phase 2 runs). **Claude** also changed significantly on all three metrics (Bonferroni-adjusted p = 1.4 × 10⁻⁵ for MDPE and MDAPE, p = 1.5 × 10⁻⁴ for Wobble), but the direction of change for MDAPE was upward at the median, from 0.0048 % in Phase 1 to 0.0079 % in Phase 2. Rather than interpreting this as a loss of accuracy, we note that the Phase 2 constraint (classical RK4 at dt = 1 s) produced a narrowing of the distribution around a systematic sub-percent bias common to all LLMs at this step size, whereas Claude’s Phase 1 distribution — which was already RK4-based — occasionally included runs that happened to sit closer to the reference than the RK4/dt = 1 s mode. Significant does not mean “improved” for Claude on MDAPE, and we avoid that phrasing. **DeepSeek** and **Grok** showed modest Phase 1-to-Phase 2 shifts on MDAPE (Bonferroni-adjusted p = 0.036 and 0.035, respectively), reaching α = 0.05 but not α = 0.01. For Wobble, the DeepSeek comparison reached α = 0.05 (p = 0.036), whereas the Grok comparison did not (p = 0.056). These shifts were driven primarily by elimination of the Phase 1 Euler-based outliers rather than a central-tendency shift. **Gemini** showed no significant Phase-to-Phase shift on any metric after Bonferroni correction (adjusted p = 0.45 for MDPE; p = 1.00 for MDAPE and Wobble), because its Phase 1 distribution was already tightly concentrated near the reference, in part because six of the twenty Gemini Phase 1 runs had self-selected sub-second step sizes.

Thus, enforcing RK4 with a fixed step size had its clearest and most strongly significant effect on ChatGPT, which had defaulted to Euler integration in Phase 1. Claude also showed a statistically significant but directionally different shift despite already being RK4-based in Phase 1, reflecting distribution narrowing rather than improvement. DeepSeek and Grok showed weaker but α = 0.05–level shifts driven by elimination of their Phase 1 Euler-based outliers, and Gemini showed no detectable improvement because its Phase 1 distribution was already tightly concentrated near the reference (in part because six of its twenty Phase 1 runs had self-selected sub-second step sizes).

### 3.5. Clinical classification of runs

To translate the metric-level analysis into a clinically interpretable outcome, each of the 200 runs was classified by MDAPE into four ordinal categories (Figure 2). Across all 200 runs: Class A (MDAPE < 1 %, excellent numerical agreement): 195 runs (97.5 %, 95 % CI 94.3–98.9 %); Class B (1 % ≤ MDAPE < 30 %, Varvel-acceptable): 1 run (0.5 %); Class C (MDAPE ≥ 30 %, clinically unacceptable): 3 runs (1.5 %); Class D (execution failure): 1 run (0.5 %). Combining all non–Class-A outcomes yields an observed overall failure rate of 5/200 (2.5 %, 95 % CI 1.1–5.7 %).

ChatGPT and Claude produced 40/40 Class A runs across both phases, with zero runs in any other category. Gemini produced 39/40 Class A runs plus the single Class D execution failure. DeepSeek produced 38/40 Class A runs, one Class B (Phase 2 run 14, MDAPE = 12.9 %) and one Class C (Phase 1 run 05, MDAPE = 99,762 %, see below). Grok produced 38/40 Class A runs plus two Class C runs in Phase 2 (runs 13 and 18, MDAPE = 97.9 % and 51.8 %, respectively).

The five non–Class-A outcomes — runs in which the generated code either failed outright, diverged beyond the Varvel acceptance threshold, or remained within that threshold but contained a structural implementation error — are examined individually in the next section. Throughout the remainder of the paper we adopt two complementary definitions of “failure”:

1. **Clinically unacceptable numerical output** (Class C + Class D): a run that produced either a plasma concentration trajectory exceeding the Varvel threshold of MDAPE ≥ 30 % or no usable trajectory at all. In our dataset this rate is 4/200 (2.0 %, 95 % CI 0.8–5.0 %).
2. **Structural-defect rate** (all runs whose failure mechanism traced to a discrete implementation bug — unit/time-scale mismatch, duplicated-bolus handling, or malformed f-string formatting): 5/200 (2.5 %, 95 % CI 1.1–5.7 %). This set is a strict superset of definition (1) because it additionally includes one Class B run (DeepSeek Phase 2 run 14, MDAPE 12.9 %) whose numerical error was within Varvel acceptance but whose cause — duplicate bolus delivery — is clearly a structural bug that would be unsafe in any clinical extrapolation.

The distinction matters because the rank-based central-tendency statistics (§2.7) are insensitive to the distinction between a Class B Varvel-acceptable MDAPE produced by a structural bug and a Class B MDAPE produced by mild numerical drift; Class B/C/D classification is the appropriate outcome for the clinical-safety component of the evaluation, while the statistical tests characterize central-tendency differences between LLMs.

### 3.6. Implementation-level analysis of divergent runs

Programmatic parsing of the 200 generated Python files recovered the internal integration algorithm and step size for every run. The distribution is summarized in Tables 1 and 2 and shown in Figure 3A,B.

**Table 1.** Algorithm and step-size choices by LLM in Phase 1 (integrator free). Summary of the integration algorithm and internal step size extracted from each of the 20 Phase 1 runs per LLM. Algorithms are listed with their frequency of use; step sizes are given in seconds after unit normalization.

| LLM | n | Algorithm | Step size (seconds) |
| --- | --- | --- | --- |
| ChatGPT | 20 | Euler: 20 | 1.0 s: 20 |
| Claude | 20 | RK4: 20 | 1.0 s: 20 |
| DeepSeek | 20 | Euler: 17, RK4: 3 | 1.0 s: 19, 6.0 s: 1 |
| Gemini | 20 | Euler: 18, RK4: 2 | 1.0 s: 14, 0.1 s: 5, 0.01 s: 1 |
| Grok | 20 | Euler: 20 | 1.0 s: 20 |

**Table 2.**
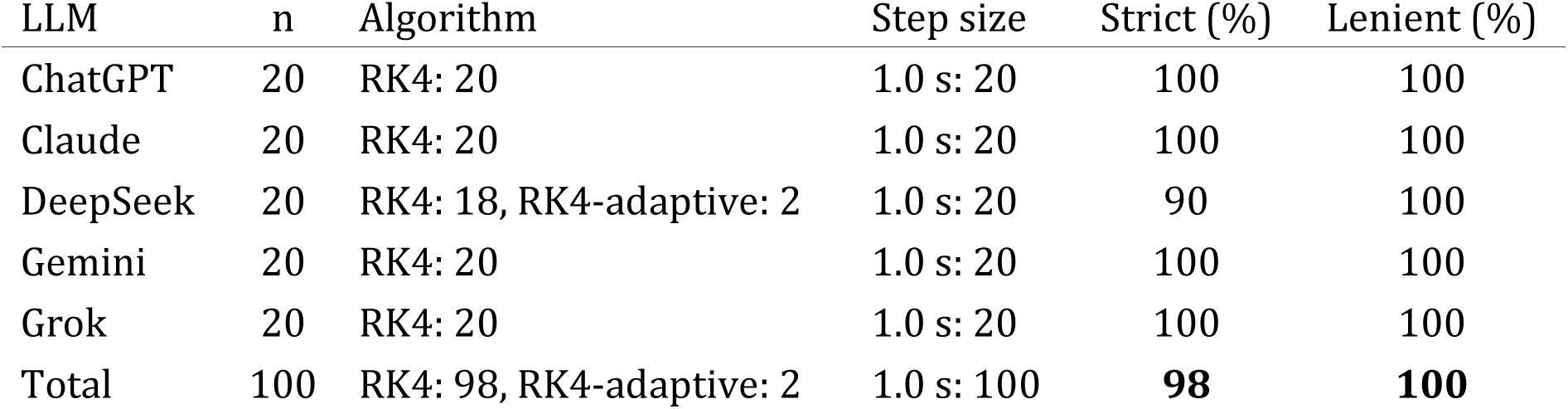
Compliance with Phase 2 prompt directives (RK4 with dt = 1 s). Summary of integration algorithm and internal step size extracted from each of the 20 Phase 2 runs per LLM. The “RK4-family (%)” column reports the lenient compliance rate that counts both classical RK4 and the RK4-adaptive variant as compliant; the strict classical-RK4 column is shown alongside. The two DeepSeek runs that used “RK4 with adaptive step-size correction” are scored as compliant under the lenient definition (RK4 superset accepted) but not under the strict definition (classical RK4 only). Per-LLM strict and lenient compliance rates with full denominators are tabulated in Supplementary Table S1b.

| LLM | n | Algorithm | Step size | Strict (%) | Lenient (%) |
| --- | --- | --- | --- | --- | --- |
| ChatGPT | 20 | RK4: 20 | 1.0 s: 20 | 100 | 100 |
| Claude | 20 | RK4: 20 | 1.0 s: 20 | 100 | 100 |
| DeepSeek | 20 | RK4: 18, RK4-adaptive: 2 | 1.0 s: 20 | 90 | 100 |
| Gemini | 20 | RK4: 20 | 1.0 s: 20 | 100 | 100 |
| Grok | 20 | RK4: 20 | 1.0 s: 20 | 100 | 100 |
| Total | 100 | RK4: 98, RK4-adaptive: 2 | 1.0 s: 100 | **98** | **100** |

In Phase 1, when integrator choice was free, the five LLMs differed sharply both in their default choice of integration algorithm and in the internal step size they selected. ChatGPT and Grok used the explicit (forward) Euler method in 20/20 Phase 1 runs; Claude used RK4 in 20/20 runs; DeepSeek used Euler in 17/20 runs and RK4 in 3/20; and Gemini used Euler in 18/20 runs and RK4 in 2/20. With respect to step size, ChatGPT, Claude, and Grok selected an effective step of 1 s in all 20 runs (written either directly as `dt` `=` `1` in seconds or as the equivalent `dt_min` `=` `1/60` in minutes); DeepSeek did so in 19/20 runs, with its single outlier (run 05) corresponding to an effective 6 s step produced by a unit misinterpretation (see below). Gemini was the only LLM to self-select sub-second step sizes, using dt = 1 s in 14/20 runs, dt = 0.1 s in 5/20 runs, and dt = 0.01 s in 1/20 runs — i.e., substantially finer than the 1-second output sampling required. In Phase 2, 98/100 runs implemented the prompt directives exactly as specified (strict compliance 98 %, 95 % CI 93.1–99.5 %), while the remaining two runs (both from DeepSeek) implemented “RK4 with adaptive step-size correction,” a superset of classical RK4; under a lenient compliance definition that accepts RK4-superset implementations, 100/100 runs were compliant (100 %, 95 % CI 96.3–100 %). All 100 Phase 2 runs used dt = 1 s as required. Both compliance rates are reported because the strict rate reflects literal adherence to the prompt wording, whereas the lenient rate reflects functional adherence to the intent of the directive.

The relationship between step size and MDAPE in Phase 1 is instructive. Among the Gemini runs, the six with sub-second step sizes achieved the lowest per-run MDAPE values observed anywhere in the dataset (median 0.0024 % at dt = 0.1 s and 0.0016 % at dt = 0.01 s, versus a median of 0.017 % at dt = 1 s across all LLMs), confirming that step-size refinement produced measurable — if clinically negligible — gains in numerical accuracy. Crucially, however, the single Class C run in Phase 1 (DeepSeek `p1_run05`, MDAPE = 99,762 %) corresponded to an **effective integration step of 6 s**, not a shorter one: inspection of the source revealed that the run declared `dt` `=` `0.1 # integration step size in minutes` while simultaneously stating “Euler method with 0.1 sec steps” in its module docstring — a unit mismatch between the author’s stated intent (0.1 s) and the actual time scale used in the Euler update (0.1 min = 6 s). Because the code integrated at effective dt = 6 s with an explicit Euler scheme, well beyond the stability limit of the three-compartment PK system, the solution diverged catastrophically.

The three numerically divergent Phase 2 runs (DeepSeek `p2_run14`, Grok `p2_run13`, Grok `p2_run18`) all declared dt = 1 s and RK4, consistent with the Phase 2 directive. Inspection of these sources revealed three implementation defects falling into two distinct error classes, neither of which is related to integrator choice:

- **Grok `p2_run13`** and **Grok `p2_run18`**: both runs retained the Marsh micro-rate constants in their native units of min⁻¹ but integrated with dt expressed in seconds, without applying the required unit conversion (division by 60). In both cases, the per-second application of a per-minute decay constant inflated total clearance by a factor of 60, driving plasma concentrations toward zero much faster than the reference and producing large negative MDPE values with correspondingly large MDAPE (Grok `p2_run13`: MDAPE = 97.9 %; Grok `p2_run18`: MDAPE = 51.8 %).
- **DeepSeek `p2_run14`**: the 140 mg bolus was effectively applied twice — once as the initial condition A_1_(0) = 140 mg before the integration loop, and again on

the first iteration of the loop, because the run’s dosing-schedule function returned a combined `bolus` + `infusion_rate` at t = 0. The double bolus plus a subtly mis-aligned early infusion schedule produced a persistent positive bias in plasma concentration (MDPE = +12.9 %, MDAPE = 12.9 %). Notably, the study prompt specified both the bolus and the first infusion interval (`t` `=` `0-10 min: continuous infusion at 700 mg/h`) as co-occurring at t = 0, mirroring an ambiguity that exists in most real-world clinical-simulator interfaces (including the Tivatrainer UI constraint described in §2.3). The Phase 2 run 14 failure suggests that at least some LLMs treat this temporal coincidence as a single combined event rather than as two separate events, motivating prompt authors to specify bolus and infusion start times explicitly and non-coincidentally.

The Class D run, Gemini `p2_run18`, presents a subtler case. The generated Python script started, imported its modules, opened a CSV file for writing, and wrote the header line `time_sec,Cp`; inside the subsequent write loop, however, the LLM emitted an f-string format specifier with an inadvertent whitespace (f“{row[1]:.6 f}”), which at Python formatting time raised `ValueError: Invalid format specifier ‘.6 f’ for object of type ‘float’` and terminated the script before any data rows were written. The resulting CSV contains only the header line and no data, so all MDPE/MDAPE/Wobble metrics are undefined for this run. From an end-user perspective this is a silent failure: although the interpreter did raise a Python-level exception, the exception occurred inside a simulation loop far from the most visible outputs of the script, and an output file exists on disk with a valid header. Without explicit inspection of either the file length or the stderr stream an operator might not notice that no simulation output was produced. We classify this run as Class D (execution failure resulting in no usable output) rather than as a Class C numerical divergence.

### 3.7. Summary of findings

Taken together, these results show three consistent patterns. First, **the overwhelming majority of LLM-generated Marsh-model code runs (97.5 %, 95 % CI 94.3–98.9 %) produced numerical output indistinguishable from a validated reference implementation at any clinically meaningful level of accuracy** (MDAPE < 1 %). Second, **two small but non-zero subsets of runs were identified: 4/200 runs (2.0 %, 95 % CI 0.8–5.0 %) produced clinically unacceptable numerical output or no usable output at all (Class C and Class D combined), and 5/200 runs (2.5 %, 95 % CI 1.1–5.7 %) exhibited structural implementation defects of any severity** — the latter set additionally including one Class B run (DeepSeek Phase 2 run 14) whose MDAPE was within Varvel acceptance but whose cause was a duplicate-bolus handling bug. Every one of these defective runs traced back to a discrete implementation error rather than to the choice of integrator itself: three of the five structurally defective runs (DeepSeek Phase 1 run 05, Grok Phase 2 runs 13 and 18) were caused by inconsistent handling of time units (minutes versus seconds), one (DeepSeek Phase 2 run 14) by a duplicated bolus administration, and one (Gemini Phase 2 run 18) by a Python-level f-string formatting error that silently truncated the output. Third, **explicit directives in the prompt (RK4, dt = 1 s) were followed at 98 % strict and 100 % lenient compliance (98/100 and 100/100 runs, respectively) in Phase 2**, demonstrating that user-side specification of numerical choices is an effective mitigation for the algorithmic component of the error distribution — but does not by itself eliminate unit-level or structural bugs, which can still manifest under a fully compliant RK4/1 s configuration.

## 4. Discussion

### 4.1. Principal findings

To the best of our knowledge, this study provides the first systematic evaluation of LLM-generated, directly executable pharmacokinetic simulation code for the Marsh propofol model using clinically established TCI validation metrics (MDPE, MDAPE, Wobble) applied to the runtime numerical output of the generated code, with repeated independent trials per LLM across five contemporary LLMs from five different providers. Across 200 code-generation runs spanning five LLMs and two prompt configurations, three principal findings emerged.

First, LLM-generated code for the Marsh three-compartment propofol model is, on the whole, highly accurate. Median MDAPE across every LLM × phase cell lay between 0.0043 % and 0.020 % — three to five orders of magnitude below the conventional Varvel acceptability threshold of MDAPE < 30 % [11], and comfortably within the numerical precision expected of hand-written scientific code. Nineteen of every twenty runs produced plasma-concentration trajectories indistinguishable from the reference implementation at any clinically meaningful level. Taken on its own, this result suggests that, for the class of task used here — a fully specified fixed-parameter three-compartment PK simulation with an explicit dosing schedule and output format — contemporary LLMs can translate the specification into numerically accurate executable code on the order of 97–99 % of the time. We deliberately phrase this as a claim about the translation step from a rich, constrained specification to runnable code, not as a claim that the models have internalized propofol pharmacokinetics: the prompt supplied the PK parameters, the dosing regimen, the integration horizon, the output schema, and the column names, so the task measured is closer to specification-guided code synthesis than to pharmacological recall.

Second, despite this high overall accuracy, a small but non-zero fraction of runs (2.0 %, 4/200; 95 % CI 0.8–5.0 %) produced output that was clinically unacceptable or non-executable (Class C + D), and an additional Class B run (DeepSeek `p2_run14`) was Varvel-acceptable by MDAPE yet contained a duplicated-bolus implementation bug detectable only by source inspection — bringing the structural-defect rate to 2.5 % (5/200; 95 % CI 1.1–5.7 %) under the broader definition that includes structurally-flawed runs whose numerical error happened to fall within the Varvel threshold. Three of the four Class C/D failures were **silent numerical failures**: scripts that ran to completion without raising any error a casual user would notice, produced a well-formed output CSV, and yielded concentration trajectories whose shape was qualitatively plausible but numerically wrong because of time-scale/unit-handling errors in the integration loop. In two Grok runs (`p2_run13` and `p2_run18`), min⁻¹ rate constants were applied per second without unit conversion, inflating clearance by a factor of 60; in one DeepSeek run (`p1_run05`), a minute-as-second step-size declaration produced an effective 6-second Euler step that drove the explicit Euler integration outside its stability region, producing catastrophic numerical divergence (MDAPE ≈ 10⁵ %). The fourth (Gemini `p2_run18`) was a **superficially-successful structural failure**: it raised `ValueError` deep inside the write loop but left a header-only CSV on disk, so a user not explicitly checking file length or stderr could plausibly believe the simulation had run. None of the five defective runs were detectable from visual inspection of the concentration trajectory alone.

Third, explicit prompt directives specifying the integrator (RK4) and the step size (dt = 1 s) were followed at 98 % strict compliance (98/100 Phase 2 runs; 95 % CI 93.1–99.5 %) and 100 % lenient compliance (accepting RK4 with adaptive step-size correction as a superset of classical RK4; 100/100 runs; 95 % CI 96.3–100 %). This demonstrates that user-side specification of numerical methods is an effective mitigation for the algorithmic component of the error distribution — in Phase 1, ChatGPT and Grok defaulted to explicit Euler integration in all 20 runs, and the prompt directive reliably replaced this with RK4. However, all three numerically divergent Phase 2 runs occurred under nominally compliant RK4/dt = 1 s configurations, and the fourth non-Class-A Phase 2 outcome was the formatting failure (Gemini `p2_run18`) — showing that prompt-level algorithmic control does not eliminate unit-level or structural bugs elsewhere in the code.

### Numerical-criterion acceptability does not imply structural correctness

One run in the present dataset (DeepSeek Phase 2 run 14) achieved an MDAPE of 12.9 %, comfortably within the Varvel acceptability threshold of 30 % that has been applied to commercial TCI systems for over three decades [11]. Source inspection, however, revealed that this numerical acceptability was produced in spite of — not because of — a faithful implementation of the dosing schedule: the 140 mg bolus was administered twice, once as an initial condition A_1_(0) = 140 mg and again on the first iteration of the integration loop, because the dosing-schedule function returned `bolus` + `infusion_rate` at t = 0. The net effect on the plasma-concentration trajectory was a persistent positive bias of approximately 12.9 %, sufficient to sit within Varvel acceptance yet indisputably an implementation defect that would be unsafe to propagate into any clinical extrapolation. This run, taken in isolation, illustrates a limitation of the Varvel criterion as a sole acceptance filter for LLM-generated code: Varvel’s metrics were designed to characterize the residual inaccuracy of otherwise-correct PK software, not to detect structural bugs in newly-written code. An MDAPE-based acceptance is therefore necessary but not sufficient; it should be complemented by a structured source-level audit of at least three implementation points — bolus event handling, infusion-rate scheduling, and unit conversion between rate constants and the integration time variable — that, in our dataset, collectively account for every structural defect observed.

### 4.2. Inter-LLM heterogeneity and its implementation-level origins

Although all five LLMs clustered at sub-percent median MDAPE, significant heterogeneity was detectable in their distributions. Claude produced the lowest variance in Phase 1 and was the only LLM that selected RK4 by default in 20 out of 20 Phase 1 runs. ChatGPT and Grok defaulted to Euler in 20 out of 20 Phase 1 runs. Gemini was also predominantly Euler (18 of 20 Phase 1 runs) but was the only LLM to self-select sub-second step sizes — dt = 0.1 s in five runs and dt = 0.01 s in one run — which produced the best per-run MDAPE values observed anywhere in the dataset (median 0.0024 % and 0.0016 %, respectively). DeepSeek used a predominantly Euler default with a small RK4 minority (3 of 20). In this dataset, every Class C numerical failure (3/200) and the single Class B duplicated-bolus run (1/200) occurred in DeepSeek or Grok, whereas the single Class D execution failure (1/200) occurred in Gemini. We note this pattern without attempting to establish provider-level reliability rankings: at n = 20 runs per LLM per phase, the present study is not powered for such rankings, and the observation should be treated as hypothesis-generating for future work at larger per-LLM sample sizes.

These differences appear to reflect consistent, model-specific defaults in the statistical patterns each LLM learned during training, rather than random variation. The fact that, in Phase 1, ChatGPT selected Euler in 20/20 runs without a single RK4 default and that Claude selected RK4 in 20/20 runs without a single Euler default, is notable: each LLM had access to the same public documentation and the same Marsh-model literature, yet their “preferred” numerical methods diverge systematically. For a clinician delegating PK code generation to an unfamiliar LLM, this implies that the baseline quality of the output — before any prompt engineering — depends substantively on which model is used, in ways that are not obvious from the generated code’s surface appearance.

The observation that every Class C numerical failure occurred in DeepSeek or Grok, while the single Class D formatting failure occurred in Gemini, is consistent with, but not demonstrative of, several plausible explanations: differences in training-data coverage of pharmacokinetic reference implementations, in post-training refinement on scientific-computing tasks, or in the scale of the underlying models. The present study is not designed to distinguish among these possibilities, and per-LLM confidence intervals (0/40, 95 % CI 0.0–8.8 % for ChatGPT and Claude; 1/40, 95 % CI 0.4–12.9 % for Gemini; 2/40, 95 % CI 1.4–16.5 % for DeepSeek and Grok) overlap extensively. Any such provider-level inference should therefore be treated as hypothesis-generating, not as a definitive ranking.

### 4.3. Relationship to prior LLM-pharmacometrics studies

The results extend prior work on LLMs in pharmacometric coding [1,2,3,4,5,6] in three ways.

First, previous evaluations have relied primarily on expert code review, visual inspection of concentration–time profiles, or non-compartmental parameter comparison [1,2,3,4]; Zheng et al. complement these with a rubric-based scoring of NONMEM code along dimensions such as syntax correctness, structural fidelity, and dataset alignment [6]. None has applied the PE/MDPE/MDAPE framework used in TCI-system validation. By adopting a metric that is (i) objectively reproducible, (ii) distributional rather than point-valued, and (iii) already familiar to clinical anesthesiologists and pharmacometricians reading our results, we believe the quantitative bar for LLM-generated PK code evaluation can be raised without requiring domain-specific re-training of readers. Critically, evaluating the *runtime numerical output* of the generated code — rather than its syntactic or structural correctness — gives direct visibility into a failure mode that static code review can miss: generated programs that parse and execute without raising an error yet produce clinically unusable concentration–time trajectories, or (as in one run in this dataset) produce no data at all despite completing successfully.

Second, Cha et al. [3] reported that ChatGPT successfully simulated a three-compartment meropenem model with a parameter difference of less than 1 % versus NONMEM, but struggled with a more complex two-compartment nivolumab model that involved multiple covariates; Zheng et al. [6] independently observed the same pattern in a rubric-based evaluation of 13 NONMEM tasks, with indirect-response and absorption-lag models yielding lower baseline scores than simple IV models. Our results corroborate the benign end of this spectrum for a structurally similar three-compartment model (< 0.02 % median MDAPE for ChatGPT on Marsh in both phases), and are consistent with the hypothesis that LLM performance degrades with model complexity. The second, more complex portion of that hypothesis — whether the failure rate rises sharply for Schnider and Eleveld propofol models with their covariate-dependent parameters and, in the case of Eleveld, non-trivial allometric scaling — is directly testable in the planned follow-up phases of the present project and is the subject of ongoing data collection.

Third, and perhaps most relevant to clinical safety, Shin et al. [1], Cloesmeijer et al. [2], and the recent synthesis by Tosca et al. [12] all report that LLM-generated pharmacometric code contains subtle errors, hallucinations, or reproducibility issues that require expert revision; Tosca et al. specifically identify the under-representation of pharmacometric data in general-purpose LLM training corpora as a fundamental limit on current applicability [12]. Our data quantify these observations precisely, but with an important caveat regarding generalization. Within the sampled mixture of five LLMs, the pooled per-run probability of a clinically unacceptable or unusable output was 2.0 % (4/200; 95 % CI 0.8–5.0 %), and the broader structural-defect or non-Class-A rate was 2.5 % (5/200; 95 % CI 1.1–5.7 %), with an expected ∼22 % cumulative probability of at least one non-Class-A or structurally defective output over 10 independent simulations (1 − (1 − 0.025)¹⁰ ≈ 0.224) when a random mixture of the five models is used. Critically, this pooled rate masks substantial LLM-level heterogeneity: the failure count was 0/40 for both ChatGPT and Claude (per-LLM failure rate 0 %, 95 % CI 0.0–8.8 %), 1/40 for Gemini (2.5 %, 95 % CI 0.4–12.9 %), 2/40 for DeepSeek (5.0 %, 95 % CI 1.4–16.5 %), and 2/40 for Grok (5.0 %, 95 % CI 1.4–16.5 %). These per-LLM confidence intervals overlap extensively, and the observed ordering must therefore be interpreted as exploratory. What the data can be said to show is that the pooled rate of 2.5 % is a mixture distribution over LLMs that clearly differ in their failure behavior; the pooled figure should accordingly not be read as a general property of “current LLMs.” For any workflow in which generated code is used to inform dosing decisions, reference-based validation should be treated as a universal requirement rather than as contingent on which specific LLM happens to be used.

### 4.4. Clinical implications

Taken together, these findings support three concrete guidelines for clinicians and researchers who use LLM-generated code in pharmacokinetic workflows.

1. **Specify the numerical method explicitly.** The 98 % strict and 100 % lenient compliance rates with RK4 and dt = 1 s directives in Phase 2 (98/100 and 100/100 runs respectively), combined with the ChatGPT / Grok habit of defaulting to Euler in Phase 1, argue strongly for including algorithm and step-size constraints in the prompt whenever accuracy matters. This is a zero-cost intervention and measurably shifts the distribution of generated code toward better numerical methods.
2. **Always validate against a reference.** Prompt directives do not prevent unit-mismatch or bookkeeping bugs. In our dataset, all three numerically divergent Phase 2 runs occurred under full RK4 compliance, and the single Class B run (duplicated-bolus bug) was Varvel-acceptable despite its structural defect. Reference-based validation is the only reliable detection mechanism for this class of failure.
3. **Do not substitute LLM-generated simulations for validated clinical tools.** The failure modes we observed — silent 60-fold clearance errors, silent duplicated-bolus administration — would, if translated uncritically from simulation to a dosing recommendation, be expected to result in unsafe clinical decisions. LLM-generated code should be regarded as a productivity aid for exploratory and educational work, not as a replacement for regulated pharmacokinetic software.

#### Operationalizing reference-based validation: a minimum validation triplet

The practical objection to “validate against a reference” is that clinicians do not typically maintain a validated reference simulator on hand. We therefore propose a minimum validation triplet, comprising three scalar comparisons that a user can compute in under a minute given any validated clinical TCI tool (e.g., Tivatrainer) or a pre-validated reference CSV such as the one released with this paper:

1. peak plasma concentration immediately after bolus administration (i.e., C_p_ at t ≈ 1 s under a 70 kg 140 mg bolus);
2. plasma concentration at end-of-infusion (C_p_ at t = 3600 s under the Marsh standard regimen);
3. plasma concentration at simulation horizon (C_p_ at t = 7200 s under the Marsh standard regimen).

Agreement within 1 % on all three of these points is sufficient to rule out every structural failure mode observed in the present dataset: a 60-fold clearance inflation driven by unit mismatch (Grok Phase 2 runs 13 and 18) and a catastrophically divergent Euler integration (DeepSeek Phase 1 run 05) both produce gross deviation at (i)–(iii); a duplicated-bolus bug (DeepSeek Phase 2 run 14) approximately doubles the initial plasma concentration and is therefore caught at (i) with a ∼100 % deviation; and a file-truncation failure (Gemini Phase 2 run 18) produces missing values at all three. This triplet therefore provides end-users with an operational, zero-cost safety check that is directly actionable in clinical and research environments where LLM-generated pharmacokinetic code is being considered. Every bug we identified is of a kind that reference-based validation at these three points would have detected; the triplet constitutes the minimum operational safeguard required for routine use of LLM-generated PK code in any setting where its numerical output might inform a clinical or research decision.

We emphasize that the scope of this triplet is deliberately narrow: it is a structural-bug screen, not a clinical-endpoint validator. The three comparison points are plasma concentration (C_p_) values because the Marsh model as originally published [7] has no effect-site compartment, and because all observed structural failures (time-scale/unit-handling errors, duplicated bolus, malformed f-string formatting) manifest as gross, ke0-independent deviations in C_p_ itself. In anesthesia practice, induction and maintenance targets are conventionally specified in terms of effect-site concentration (C_e_, e.g., C_e_ ≈ 3 µg·mL⁻¹ for propofol induction of general anesthesia), and clinical TCI systems pair each PK model with a ke0 specific to that model and usage context. Extending the present benchmark to C_e_-based validation, to the covariate-dependent Schnider [8] and allometrically-scaled Eleveld [9] models, and to clinically meaningful dosing endpoints (target achievement, time-to-target, dose adjustment under patient-specific covariates) is planned future work in this series. Visual Cp(t) and Ce(t) trajectories are already fully reproducible from the open dataset release (see Data Availability), and interactive clinical-profile visualization is envisioned as a companion tool once the Schnider/Eleveld Phase-2 data collection completes.

### 4.5. Limitations

This study has several limitations that qualify the conclusions above.

First, we evaluated a single, structurally simple PK model. The Marsh model has fixed parameters, no covariates, and no allometric scaling; the effect-site compartment (k_e0_) was not included. Models with increasing complexity — Schnider, with its weight- and age-dependent volumes; Eleveld, with population-level allometry and a richer covariate structure — may exhibit qualitatively different failure modes, and the present results should not be extrapolated uncritically. Evaluation of these models is in progress and will be reported separately.

Second, we evaluated each LLM through its public chat interface at a specific point in time, using a specific model version per LLM (documented in Supplementary Table S1). For every LLM we deliberately used the default, non-reasoning / non-thinking response mode (GPT-5.3 Instant, Claude Sonnet 4.6, DeepSeek-V3.2 in Instant mode, Gemini 3 Fast, Grok 4.1 Fast) rather than the slower, more deliberative “Thinking” / “Reasoning” variants that each provider also makes available. This choice was intentional: it reflects how a busy clinician is most likely to interact with these tools in practice — typing a question into the default interface and accepting the first response returned — and therefore provides a realistic estimate of the quality of code that non-expert users will encounter in routine use. The likely improvement achievable by switching to a reasoning variant is an important open question; because reasoning variants incur substantially higher latency and cost (and, for DeepSeek-V3.2-Speciale, are explicitly restricted to deep-reasoning tasks without tool use), characterizing that trade-off separately, rather than pooling it into a single headline failure rate, seems to us the more useful framing. LLM capabilities evolve on a timescale of weeks, and results from any given model version may not generalize. Furthermore, data collection for each LLM was staggered across several days (Supplementary Table S1b), so the runs for a given LLM represent time-windowed operational samples rather than strictly i.i.d. repeated draws; this is a mild interpretive caveat rather than a methodological defect, but readers should be aware that the behavior observed for one LLM reflects the state of that service over its specific two-day collection window. We have attempted to mitigate the overall snapshot nature of these results by archiving exact prompts, model versions, timestamps, and source-code SHA-256 hashes for every run, but the absolute failure rates we report should be treated as a snapshot rather than a permanent property of these systems.

Third, 20 runs per cell is a modest sample. For the per-LLM per-phase failure-rate confidence intervals, 20 runs provides roughly ±10 % resolution at a 5 % failure rate — adequate to rank LLMs by order-of-magnitude reliability, but insufficient to compare two LLMs with, say, 2 % and 5 % failure rates. The sample size was chosen to balance the cost of manual run execution and review against statistical power for the main comparisons; future work with automated run pipelines should aim for n ≥ 100 per cell.

Fourth, our prompt specified a single patient weight, dose, and infusion schedule. The failure modes we observed (unit mismatch, double bolus) could in principle interact non-trivially with different dosing regimens. A formal perturbation study varying these inputs would strengthen the generalizability of the failure-rate estimates.

Fifth, all runs were executed in Python 3. LLM-generated code in other languages (R, Julia, MATLAB) or in domain-specific platforms (NONMEM, Monolix, nlmixr) may behave differently. Prior work [1,3] has specifically found LLM difficulty with NONMEM control streams, suggesting the failure distribution may be substantially less favorable in those environments.

Sixth, we did not attempt prompt-level error-correction or iterative refinement. In practice, many end users would inspect the generated code, request corrections, or paste error traces back to the LLM — an interaction loop which has been shown elsewhere to substantially improve output quality [13]. Our single-shot failure rate of 2.5 % should therefore be understood as an upper bound on the rate that would persist after one round of human review.

Seventh, a methodological consideration concerns the inherent properties of rank-based non-parametric statistics. The Kruskal–Wallis, Dunn, and Mann–Whitney tests we use operate on ranks, so a catastrophically divergent run with MDAPE on the order of 10⁵ % and a mildly elevated run with MDAPE of a few percent are treated as equally “high-ranked” for the purpose of the test statistic. This is a known property of rank-based methods and is appropriate for comparing the central tendency of heavy-tailed distributions, but it means that the magnitude of clinically dangerous divergences is not directly reflected in the p-values we report. For that reason, we supplement the rank-based comparisons with Varvel-style ordinal classification (Class A/B/C/D in Figure 2) and with explicit reporting of per-run maximum absolute prediction error (§3.2); together these preserve the clinically relevant magnitude information that the rank tests abstract away.

Finally, because only one pharmacokinetic model, one dosing regimen, one programming language, and one single-turn prompt family were evaluated, the present study should be interpreted as a benchmark of a deliberately simple positive-control task rather than as a comprehensive estimate of the reliability of LLM-generated pharmacokinetic code in general. Evaluation of the covariate-dependent Schnider and allometrically-scaled Eleveld propofol models, of effect-site kinetics, of multi-turn error-correction workflows, and of non-Python coding environments, will be reported in subsequent work in this series.

### 4.6. Future directions

The present work is the first of a planned series. Immediate next steps are (i) extension to the Schnider and Eleveld models to test whether the failure rate scales with model complexity as predicted by Cha et al. [3] and independently supported by Zheng et al. [6]; (ii) extension to effect-site kinetics (k_e0_) and to target-controlled infusion simulation modes; (iii) expansion to a broader set of LLMs, including smaller open-weight models for which local-deployment reproducibility is straightforward; and (iv) evaluation of multi-turn and agentic workflows — in which an LLM generates code, inspects its own output, and revises — to quantify the error-correction benefit of a minimally structured human-in-the-loop interaction, complementing the broader agentic-workflow framework recently proposed by Shahin et al. for quantitative clinical pharmacology and translational sciences [14].

Beyond within-anesthesiology applications, this framework — a shared, standardized prompt applied to multiple LLMs, a validated reference simulator, and clinically interpretable accuracy metrics — can be transplanted to other quantitative biomedical domains where clinicians or investigators are beginning to delegate scientific-computing tasks to LLMs: PBPK modeling, ECG and hemodynamic simulation, ventilator waveform analysis, and radiation dose calculation, among others. The per-domain value is not the absolute MDAPE numbers, but the demonstration that a small, well-defined evaluation harness can distinguish clinically safe from unsafe generated code with far greater precision than expert eyeballing alone.

### 4.7. Conclusion

Contemporary large language models can generate executable Python code for the Marsh three-compartment propofol pharmacokinetic model with, in most runs, sub-percent median absolute prediction error relative to a validated reference implementation. Across 200 runs spanning five LLMs, 97.5 % (95 % CI 94.3–98.9 %) produced numerically excellent output (MDAPE < 1 %), and 99.5 % of scripts produced a valid time–concentration CSV. However, 4/200 runs (2.0 %, 95 % CI 0.8–5.0 %) produced clinically unacceptable or non-executable output (Class C + D), and an additional Varvel-acceptable run (Class B, MDAPE 12.9 %) nonetheless contained a duplicated-bolus implementation bug — illustrating that numerical acceptance alone does not imply structural correctness. Every observed failure traced to one of three discrete implementation error classes — time-scale/unit-handling errors, duplicated bolus, or malformed f-string formatting — none of which the LLMs’ own output flagged as suspect. Two further Phase 2 runs silently substituted an RK4-adaptive variant for the prompt-specified classical RK4; these were numerically benign (Class A, MDAPE < 0.01 %) but constitute strict directive non-compliance and are best categorized as prompt-adherence violations rather than as numerical failures.

These findings suggest three constructive directions. First, at the user level, the minimum reference-based validation triplet proposed in §4.4 offers a zero-cost, clinically actionable guardrail that would have detected every failure in the present dataset in under a minute of work. Second, at the prompt-engineering level, explicit specification of the numerical method and of unit-handling conventions eliminated the algorithmic component of the error distribution at ≥ 98 % strict compliance, confirming that well-designed prompts substantively shift the distribution of generated code toward safer output. Third, at the tooling level, the observed failure classes suggest concrete primitives that future LLM-assisted scientific-computing environments could incorporate — for example, automatic unit-consistency checking at the integration-loop level, automatic generation of reference-trajectory test cases from a model specification, and agentic workflows in which a second LLM or a lightweight static analyzer audits the first LLM’s output before it is handed to the user. Each of these primitives would be cheap to implement and would materially reduce the residual failure rate.

In the context of the Marsh model evaluated here, LLM-generated pharmacokinetic code is a powerful productivity tool for exploratory and educational work. At current failure rates, and given the silent or superficially-successful nature of the failure modes, it should not be substituted for validated, regulated pharmacokinetic software in clinical or safety-relevant use without independent reference-based validation. Establishing that validation as a routine, low-friction step — rather than as an optional safeguard — is the near-term path toward making LLM-assisted PK coding safe for routine biomedical use.

## Supporting information

manuscript_medRxiv.pdf

## Data Availability

The full run-level dataset (200 Python source files, 200 output CSVs, 200 per-run PE
series, and associated metadata including model version, timestamp, and SHA-256
hashes), together with the reference implementation and all analysis scripts, are re
leased under the MIT license at https://github.com/omote-masahito/llm-pk-marsh-benchmark

## Declarations

### Funding

This work received no external funding.

### Competing interests

The author declares no competing interests. Claude (Anthropic PBC) — one of the LLMs evaluated in this study — was also used as an AI assistant during code review and manuscript drafting (see Section 2.8 and the AI-assistance disclosure below). The author declares no financial relationship with Anthropic PBC, OpenAI, Google, xAI, or DeepSeek beyond paid end-user subscriptions to the respective services.

### Ethics approval

Because this study consisted entirely of in silico simulations and did not involve human subjects, animals, or identifiable patient data, institutional review board approval was not required.

### Consent to participate

Not applicable.

### Consent for publication

Not applicable.

### Data availability

The full run-level dataset (200 Python source files, 200 output CSVs, 200 per-run PE series, and associated metadata including model version, timestamp, and SHA-256 hashes), together with the reference implementation and all analysis scripts, are released under the MIT license at https://github.com/omote-masahito/llm-pk-marsh-benchmark.

### Code availability

All analysis code, figure-generation scripts, and the reference simulator are released under the MIT license at the repository indicated above.

### Author contributions

As the sole author, **Masahito Omote** was solely responsible for conceptualization, methodology, software, validation, formal analysis, investigation, data curation, writing — original draft, writing — review and editing, and visualization. The author used Claude (Anthropic PBC) as an AI assistant for portions of the code review, data analysis workflow, and manuscript drafting, with all outputs verified and approved by the author (see Section 2.8 and Acknowledgment of AI assistance below).

## Acknowledgment of AI assistance

The author used Claude (Anthropic PBC) to assist with code review, data analysis workflow, and manuscript drafting. All scientific claims, numerical results, and the final text were verified and approved by the author, who takes full responsibility for the integrity of the work. This use is disclosed in accordance with Springer Nature AI-assistance policy.

---

**Supplementary Figure S1.**
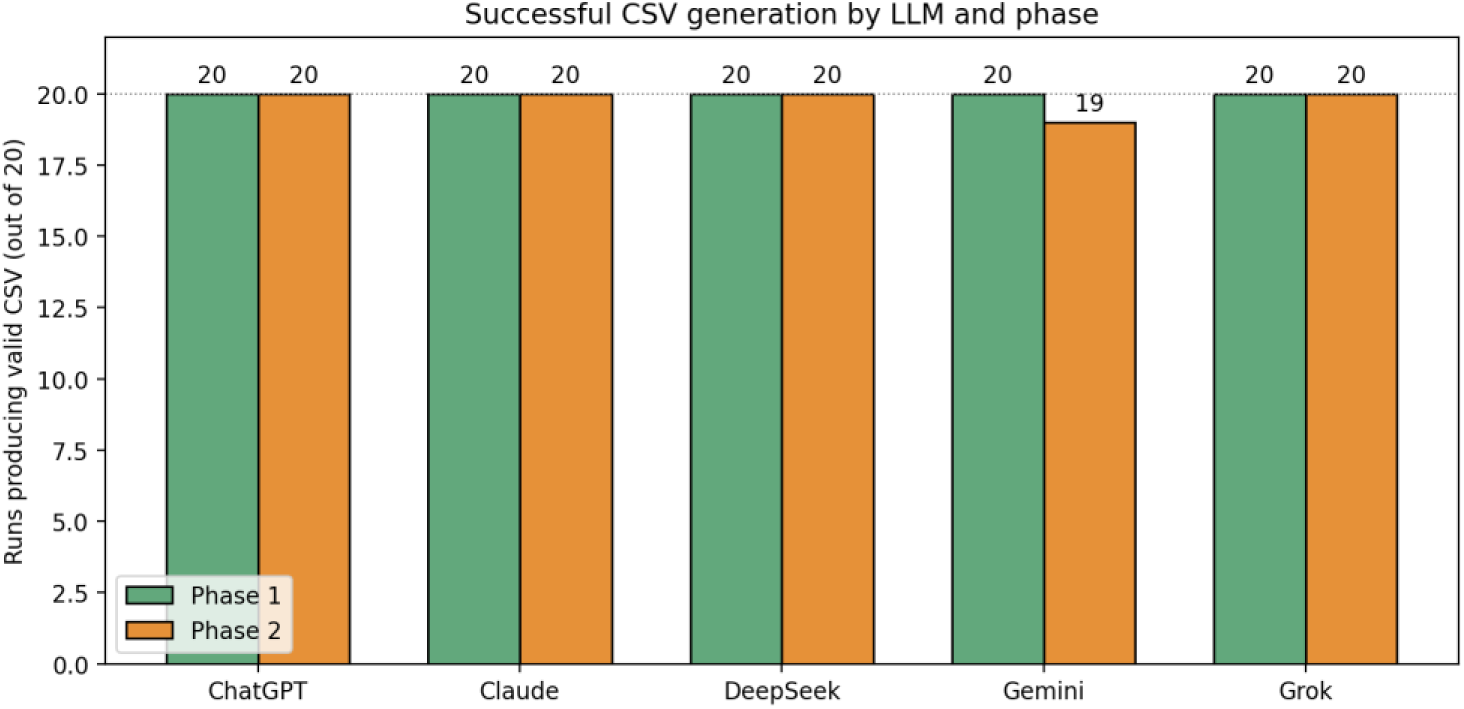
Code execution success rate across LLMs and phases. Grouped bar plot showing the proportion of 20 attempted runs per LLM per phase that produced a valid time–concentration CSV. All five LLMs achieved 20/20 success in both phases except Gemini in Phase 2, which achieved 19/20: the Phase 2 run 18 script raised `ValueError: Invalid format specifier ‘.6 f’ for object of type ‘float’` during the write loop because the LLM emitted a malformed f-string format specifier (f“{row[1]:.6 f}” with a stray whitespace), aborting the script after the CSV header row had been written and leaving the data file empty.

**Supplementary Figure S2.**
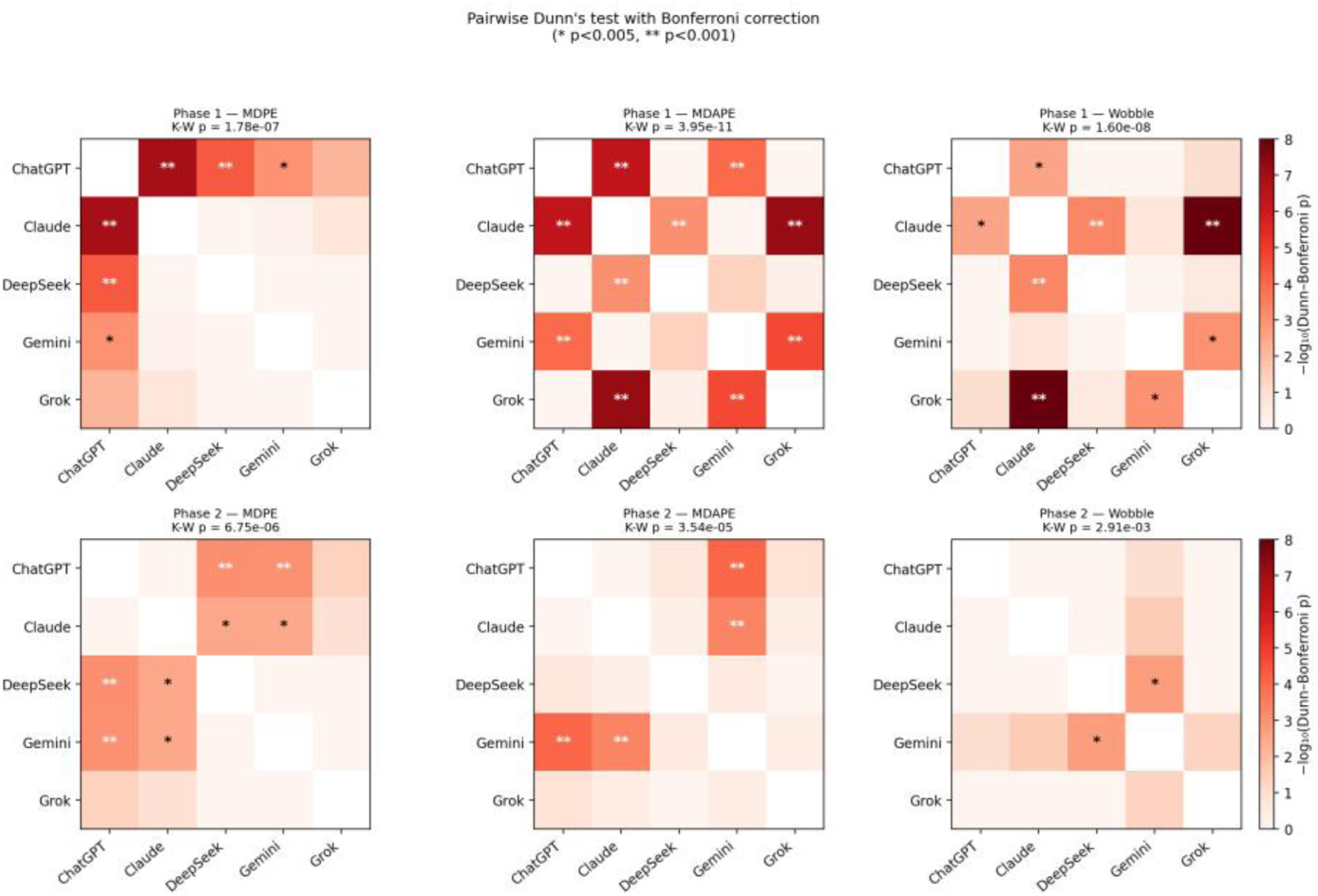
Pairwise Dunn’s test (Bonferroni-corrected) p-values for LLM-versus-LLM comparisons. Heatmap representation of all pairwise post-hoc comparisons between the five LLMs, computed using Dunn’s test with Bonferroni correction for the 10 pairwise comparisons per phase × metric cell. Results are shown separately for each metric (columns: MDPE, MDAPE, Wobble) and each phase (rows: Phase 1, Phase 2). Each 5 × 5 matrix compares one LLM against every other; the diagonal (self-comparison) is left blank. Cells are colored by −log₁₀(adjusted p), and significance markers are overlaid against the Bonferroni-adjusted p-values: * = p_adj_ < 0.05, ** = p_adj_ < 0.01. The Kruskal–Wallis omnibus p-value for each phase × metric cell is shown above the matrix. The Phase 2 Wobble panel illustrates that, after Bonferroni correction, Phase 2 inter-LLM differences are primarily MDAPE-dominated rather than Wobble-dominated.

## Notes

### Competing Interest Statement

The authors have declared no competing interest.

## References

1. Shin E, Yu Y, Bies RR, Ramanathan M. Evaluation of ChatGPT and Gemini large language models for pharmacometrics with NONMEM. J Pharmacokinet Pharmacodyn. 2024;51(3):187–197.

2. Cloesmeijer ME, Janssen A, Koopman SF, Cnossen MH, Mathôt RAA; SYMPHONY consortium. ChatGPT in pharmacometrics? Potential opportunities and limitations. Br J Clin Pharmacol. 2024;90(1):360–365.

3. Cha HJ, Choe K, Shin E, et al. Leveraging large language models in pharmacometrics: evaluation of NONMEM output interpretation and simulation capabilities. J Pharmacokinet Pharmacodyn. 2025;52:34.

4. Sánchez Herrero S, Calvet L. Generative artificial intelligence models in pharmacokinetics: a study on a two-compartment population model. Research Square [preprint]. 2024. 10.21203/rs.3.rs-4693613/v1

5. Holt S, Qian Z, Liu T, Weatherall J, van der Schaar M. Data-driven discovery of dynamical systems in pharmacology using large language models. In: Proceedings of the Thirty-Eighth Annual Conference on Neural Information Processing Systems (NeurIPS 2024); 2024 Dec 10–15; Vancouver, Canada. Available from: https://openreview.net/forum?id=KIrZmlTA92

6. Zheng W, Wang W, Kirkpatrick CMJ, Landersdorfer CB, Yao H, Zhou J. AI for NONMEM coding in pharmacometrics research and education: shortcut or pitfall? CPT Pharmacometrics Syst Pharmacol. 2025;14(12):1965–1969. 10.1002/psp4.70125

7. Marsh B, White M, Morton N, Kenny GNC. Pharmacokinetic model driven infusion of propofol in children. Br J Anaesth. 1991;67(1):41–48. 10.1093/bja/67.1.41

8. Schnider TW, Minto CF, Gambus PL, Andresen C, Goodale DB, Shafer SL, Youngs EJ. The influence of method of administration and covariates on the pharmacokinetics of propofol in adult volunteers. Anesthesiology. 1998;88(5):1170–1182. 10.1097/00000542-199805000-00006

9. Eleveld DJ, Colin P, Absalom AR, Struys MMRF. Pharmacokinetic–pharmacodynamic model for propofol for broad application in anesthesia and sedation. Br J Anaesth. 2018;120(5):942–959. 10.1016/j.bja.2018.01.018

10. Gutta BV. Tivatrainer: intravenous drug simulation application. Version 2.2.0, Build 287. Aerdenhout, The Netherlands: Gutta BV; [cited 2026 Apr]. Available from: https://www.tivatrainer.com

11. Varvel JR, Donoho DL, Shafer SL. Measuring the predictive performance of computer-controlled infusion pumps. J Pharmacokinet Biopharm. 1992;20(1):63–94. 10.1007/BF01143186

12. Tosca EM, Aiello L, De Carlo A, Magni P. Pharmacometrics in the age of large language models: a vision of the future. Pharmaceutics. 2025;17(10):1274. 10.3390/pharmaceutics17101274

13. Shin E, Ramanathan M. Evaluation of prompt engineering strategies for pharmacokinetic data analysis with the ChatGPT large language model. J Pharmacokinet Pharmacodyn. 2024;51(2):101–108.

14. Shahin MH, Goswami S, Lobentanzer S, Corrigan BW. Agents for change: artificial intelligent workflows for quantitative clinical pharmacology and translational sciences. Clin Transl Sci. 2025;18(3):e70188. 10.1111/cts.70188

