## Supplementary material for "Silent numerical failures in large language model–generated pharmacokinetic simulation code: a benchmark against target-controlled infusion validation criteria using the Marsh propofol model": manuscript_medRxiv.pdf

Masahito Omote

#### Scope of this Supplementary Appendix

This Supplementary Appendix is a companion to the main manuscript. It contains the full text of the two experimental prompts, per-run metadata, the reference-implementation source and triple-validation details, extended statistical output, implementation-level failure-mode analysis, the recommended minimum validation protocol (including a standalone Python validator), and a full AI-use disclosure.

The main figures (Figures 1–4) and main tables (Tables 1–2), together with the two supplementary figures (Supplementary Figures S1 and S2), are embedded **in the main manuscript** and are not reproduced here. This appendix provides only material that does not already appear in the main manuscript: supplementary *tables* (S1 and S1b), supplementary appendices S1–S7, and supplementary references.

#### Table of Contents


1. Supplementary Appendix S1 — Full prompt text and per-run metadata
2. Supplementary Appendix S2 — Reference implementation and triple validation
3. Supplementary Appendix S3 — Extended statistical detail and within-LLM results
4. Supplementary Appendix S4 — Implementation-level failure mode analysis
5. Supplementary Appendix S5 — Recommended minimum validation protocol
6. Supplementary Appendix S6 — Extended limitations and comparison with prior work
7. Supplementary Appendix S7 — AI use disclosure
8. Supplementary Table S1 — LLM model versions, access dates, and account configurations
9. Supplementary Table S1b — Phase 2 compliance with prompt directives

10. Supplementary References

### Supplementary Appendix S1 — Full prompt text and per-run metadata

#### S1.1 Phase 1 prompt (integrator free)

The complete prompt issued to each LLM is reproduced below verbatim:

```
Please write a Python program that simulates the plasma concentration
of propofol using the Marsh three-compartment pharmacokinetic model.

Requirements:

1. Do NOT use scipy.integrate.odeint, scipy.integrate.solve_ivp, or
   any equivalent external ODE solver library. Implement the
   numerical integration from scratch.

2. Patient: 70 kg adult.

3. Marsh model parameters (original 1991 parameters):
     Vc  = 228 mL/kg
     k10 = 0.119  min^-1
     k12 = 0.112  min^-1
     k13 = 0.0419 min^-1
     k21 = 0.055  min^-1
     k31 = 0.0033 min^-1
   Reference: Marsh B, White M, Morton N, Kenny GN. Pharmacokinetic
   model driven infusion of propofol in children.
   Br J Anaesth. 1991;67(1):41-48.

4. Dosing regimen:
     - Bolus: 140 mg into the central compartment at t = 0 min
     - Continuous infusion:
         0 – 10  min :  700 mg/h
        10 – 20  min :  560 mg/h
        20 – 60  min :  420 mg/h
        60 – 120 min :    0 mg/h

5. Simulation horizon: 0 to 120 minutes.

6. Output a CSV file with two columns, "time_sec" and "Cp", where:
     - time_sec is in seconds (integer)
```

```
- Cp is the plasma concentration in µg/mL
- one row per second from t = 0 to t = 7200
```

##### S1.2 Phase 2 prompt (RK4 with dt = 1 s mandated)

The Phase 2 prompt is identical to Phase 1 except that the following sentence is inserted between requirements 1 and 2:

```
Use the classical fourth-order Runge–Kutta method (RK4) with a fixed
time step of dt = 1 second for the numerical integration.
```

##### S1.3 Notes on prompt design

The prompt was deliberately concrete about the task specification (parameters, regimen, output format) and deliberately silent about the implementation method in Phase 1. This reflects what a clinician or researcher would most plausibly write when first asking an LLM for simulation code. The Marsh parameters were transcribed from the original 1991 paper, without the covariate scaling introduced by later tuning studies.

##### S1.4 Per-run metadata

For every run, the following metadata are recorded in llm_outputs/Phase{n}/<llm>/metadata.json and released with the public dataset (see Data Availability in the main text):

- run_id, phase, timestamp (UTC)
- code_file and its code_sha256
- execution_success (boolean) and execution_error (string if applicable)
- output_csv and its output_sha256
- csv_row_count
- algorithm_used (Euler, RK4, RK4-adaptive), step_size (seconds, after unit normalization)
- vc_calculation (mL/kg × 70 = L, or direct L, or other)
- MDPE, MDAPE, Wobble, Mean_PE, SD_PE, Max_PE, n_evaluated_points
- non_integer_times, float_precision_issues, free-text notes

SHA-256 hashes enable byte-level reproducibility.

#### Supplementary Appendix S2 — Reference implementation and triple validation

A reference simulator for the Marsh model was implemented independently by the author in plain Python. The central ($V_1$ = 228 mL·kg$^{-1}$ × 70 kg = 15.96 L), peripheral, and deep peripheral compartments were modeled as a three-compartment mammillary system, with amounts $A_1$, $A_2$, $A_3$ (mg) in each compartment governed by the ODE system

$$\frac{dA_1}{dt} = I(t) - (k_{10} + k_{12} + k_{13})\, A_1 + k_{21}\, A_2 + k_{31}\, A_3$$
$$\frac{dA_2}{dt} = k_{12}\, A_1 - k_{21}\, A_2$$
$$\frac{dA_3}{dt} = k_{13}\, A_1 - k_{31}\, A_3$$

where $I(t)$ is the instantaneous infusion rate (mg·min$^{-1}$), with an instantaneous bolus added to $A_1$ at t = 0. Plasma concentration is $C_p(t) = A_1(t)/V_1$.

The reference simulator uses classical RK4 with a fixed internal time step of 0.01 s. To protect against implementation-level errors in the reference itself, the reference trajectory was validated by three independent comparisons:

1. **Self-consistency against `scipy.integrate.odeint`** under the same parameters and regimen. Maximum relative deviation across the full 0–7,200 s horizon was 0.00044 % — well below any clinically meaningful precision.

2. **Comparison against Tivatrainer (Gutta BV, version 2.2.0, build 287)** using the matching 70 kg adult, Marsh parameter set, and bolus-plus-infusion regimen. Tivatrainer exports concentrations at 1-minute intervals; at every exported time point the reference agreed to MDAPE = 0.06 % across the full horizon.

3. **Parameter verification against the original Marsh 1991 paper** [7], confirming $V_c$ = 228 mL·kg$^{-1}$ and all five micro-rate constants without the later covariate adjustments sometimes found in commercial implementations.

The reference simulator source (`reference/reference.py`), its exported CSV trajectory (`reference/reference.csv`), and the Tivatrainer export used for cross-validation (`reference/tivatrainer_Propofol_Marsh.csv`) are released with the dataset.

### Supplementary Appendix S3 — Extended statistical detail and within-LLM results

#### S3.0 Primary vs exploratory outcomes

The pre-specified primary outcome of this study is the pooled proportion of runs across all 5 LLMs and both phases that produced clinically unacceptable numerical output or execution failure (Class C + D), reported with a two-sided 95 % Wilson score confidence interval. In this dataset: **4/200 = 2.0 % (95 % CI 0.8–5.0 %)**. A secondary structural-defect rate additionally counts Class B runs with an identifiable implementation bug (DeepSeek `p2_run14`, duplicated bolus, MDAPE 12.9 %), yielding **5/200 = 2.5 % (95 % CI 1.1–5.7 %)**. Per-LLM comparisons of failure rate and of metric distribution are treated throughout as **exploratory**: at n = 20 runs per LLM per phase, the study is not powered to establish provider-level ordering of failure risk, and all between-LLM differences reported are interpreted as hypothesis-generating for future work.

We also adopt a two-term failure taxonomy that the main text uses throughout. **Silent numerical failures** are runs that completed without error, wrote a well-formed CSV, and produced a plausible-looking but numerically wrong trajectory (the three unit-mismatch Class C runs). **Superficially-successful structural failures** are runs that produced output passing a superficial existence check but with a structural defect (Gemini `p2_run18`'s header-only CSV after mid-write `ValueError`). Class B runs with identifiable structural bugs sit at the intersection; the primary outcome counts only numerical unacceptability, while the secondary structural-defect rate additionally captures these Varvel-acceptable structurally-flawed runs.

#### S3.1 Per-LLM pairwise Dunn–Bonferroni results

In Phase 1, Claude was significantly separated from ChatGPT on MDAPE ($p_{adj} = 4.7 \times 10^{-7}$), from Grok on MDAPE ($p_{adj} = 6.2 \times 10^{-8}$) and Wobble ($p_{adj} = 4.7 \times 10^{-9}$), and from DeepSeek on MDAPE ($p_{adj} = 9.3 \times 10^{-4}$) and Wobble ($p_{adj} = 5.1 \times 10^{-4}$). Gemini was significantly separated from ChatGPT on MDAPE ($p_{adj} = 1.0 \times 10^{-4}$) and from Grok on both MDAPE ($p_{adj} = 1.9 \times 10^{-5}$) and Wobble ($p_{adj} = 1.0 \times 10^{-3}$), reflecting the influence of its sub-second step-size choices. In Phase 2, Gemini was separated from ChatGPT (MDAPE $p_{adj} = 8.1 \times 10^{-5}$) and from Claude (MDAPE $p_{adj} = 4.6 \times 10^{-4}$), but these Phase 2 differences no longer reached the Bonferroni-adjusted threshold on Wobble (ChatGPT vs Gemini $p_{adj} = 9.4 \times 10^{-2}$, Claude vs Gemini $p_{adj} = 2.7 \times 10^{-2}$), indicating that the Phase 2 inter-LLM differences are MDAPE-dominated rather than Wobble-dominated.

ChatGPT and Claude did not differ significantly in Phase 2 on any metric (MDAPE $p_{adj} = 1.00$); we cannot conclude from this that the two distributions are equivalent, only that our sample was insufficient to detect a difference. Grok's Phase 2 distribution was not significantly separated from other LLMs after Bonferroni correction, despite raw Mann–Whitney p-values below 0.05 in several comparisons, consistent with two divergent runs inflating its variance without shifting its central tendency. The full pairwise p-value matrices are shown in Supplementary Figure S2.

#### S3.2 Within-LLM Phase 1 vs Phase 2 changes

**ChatGPT** showed the largest shift, with all three metrics significantly changing from Phase 1 to Phase 2 (all Bonferroni-adjusted $p = 1.2 \times 10^{-8}$). Median MDAPE decreased from 0.017 % to 0.0079 %, reflecting the replacement of Euler integration (used in 20/20 Phase 1 runs) by RK4 (20/20 Phase 2 runs).

**Claude** also changed significantly on all three metrics (Bonferroni-adjusted $p = 1.4 \times 10^{-5}$ for MDPE and MDAPE, $p = 1.5 \times 10^{-4}$ for Wobble), but the direction of change for MDAPE was upward at the median (0.0048 % → 0.0079 %). Rather than interpreting this as a loss of accuracy, we note that the Phase 2 constraint (classical RK4 at dt = 1 s) produced a narrowing of the distribution around a systematic sub-percent bias common to all LLMs at this step size, whereas Claude's Phase 1 distribution — which was already RK4-based — occasionally included runs that happened to sit closer to the reference than the RK4/dt = 1 s mode.

**DeepSeek** and **Grok** showed modest Phase 1-to-Phase 2 shifts on MDAPE (Bonferroni-adjusted $p = 0.036$ and 0.035, respectively), reaching $\alpha = 0.05$ but not $\alpha = 0.01$. For Wobble, the DeepSeek comparison reached $\alpha = 0.05$ ($p = 0.036$), whereas the Grok comparison did not ($p = 0.056$). These shifts were driven primarily by elimination of the Phase 1 Euler-based outliers rather than a central-tendency shift.

**Gemini** showed no significant Phase-to-Phase shift on any metric after Bonferroni correction (adjusted $p = 0.45$ for MDPE; $p = 1.00$ for MDAPE and Wobble), because its Phase 1 distribution was already tightly concentrated near the reference, in part because six of the twenty Gemini Phase 1 runs had self-selected sub-second step sizes.

#### S3.3 Rank-based statistics and heavy-tailed distributions

A methodological consideration concerns the inherent properties of rank-based non-parametric statistics. The Kruskal–Wallis, Dunn, and Mann–Whitney tests operate on ranks, so a catastrophically divergent run with MDAPE on the order of $10^5$ % and a mildly elevated run with MDAPE of a few percent are treated as equally "high-ranked" for the test statistic. This is a known property of rank-based methods and is appropriate for comparing the central tendency of heavy-tailed distributions, but it means that the magnitude of clinically dangerous divergences is not directly reflected in the reported p-values. We therefore supplement the rank-based comparisons with Varvel-style ordinal classification (Class A/B/C/D; Figure 2) and with per-run maximum absolute prediction error reporting; together these preserve the clinically relevant magnitude information that the rank tests abstract away.

#### Supplementary Appendix S4 — Implementation-level failure mode analysis

For every non-Class-A run, the generated source code was inspected to identify the proximate cause of the divergence. Five runs required this analysis. All five traced to discrete implementation bugs rather than to algorithmic error. Each bug class is described below with the exact root cause; the corresponding code is preserved byte-for-byte in the public dataset (`llm_outputs/Phase{1,2}/<llm>/p{n}_run{NN}.py`).

##### S4.1 Unit mismatch (3 runs)

**DeepSeek `p1_run05`** (Phase 1, MDAPE ≈ 99,762 %). The failure mechanism here is a *time-scale* mismatch rather than a *unit* mismatch on the rate constants. The generated code declared `dt = 0.1` with the comment `# integration step size in minutes` while simultaneously stating "Euler method with 0.1 sec steps" in its module docstring. The integration loop interpreted `dt` as 0.1 min, yielding an effective Euler step of 6 s — well beyond the stability limit of the three-compartment Marsh system. The solution diverged catastrophically owing to numerical instability of explicit Euler at this step size, producing the observed MDAPE of $\sim 10^5$ %. This is mechanistically distinct from the Grok `p2_run13`/`p2_run18` failures described next, in which the rate constants themselves were applied per-second instead of per-minute.

**Grok `p2_run13` and Grok `p2_run18`** (Phase 2, MDAPE = 97.9 % and 51.8 %). Both runs declared RK4 with dt = 1 s (consistent with the Phase 2 directive) but retained the Marsh rate constants in their native $\text{min}^{-1}$ units while integrating in seconds, producing the same kind of 60× clearance inflation.

##### S4.2 Duplicated-bolus handling (1 run)

**DeepSeek `p2_run14`** (Phase 2, MDAPE = 12.9 %). The 140 mg loading dose was injected *twice* — once as the initial condition for $A_1$ at t = 0, and once again inside the infusion loop on the first time step. The resulting concentration trajectory had the correct decay shape but an inflated magnitude. Because the MDAPE of 12.9 % is below the Varvel threshold of 30 %, this run is classified as Class B rather than Class C, but the underlying cause is a structural implementation bug that would be clinically unsafe in any extrapolation.

##### S4.3 Malformed f-string formatting (1 run)

**Gemini `p2_run18`** (Phase 2, Class D). The generated script opened a CSV file, wrote the header line (`time_sec,Cp`), and entered the simulation loop. On the first row-write, Python raised `ValueError: Invalid format specifier '.6 f' for object of type 'float'` because the LLM emitted an f-string with an inadvertent whitespace inside the format specifier:

```
csv_writer.writerow([row[0], f"{row[1]:.6 f}"])  # note the stray space
```

The exception terminated the script before any data row was written. The resulting CSV contains only the header and no data, so MDPE/MDAPE/Wobble are undefined for this run. From an end-user perspective, this is a silent failure: the interpreter did raise an exception, but it was raised deep inside a simulation loop while leaving a well-formed output file on disk. Without explicit inspection of file length or stderr, an operator might not notice that no simulation output was produced.

##### S4.4 Adaptive-step substitution outside prompt directives (2 runs)

**DeepSeek `p2_run17` and `p2_run19`** (Phase 2, both Class A). These runs implemented an RK4 integrator with automatic step-size correction — a superset of the classical RK4 that the Phase 2 prompt directive requested. Numerical output was excellent (MDAPE < 0.01 % for both runs). We nonetheless flag this as a structural class because it is a directive violation: under a stricter interpretation of the Phase 2 prompt, these runs are non-compliant. Under a lenient interpretation (which we also report), the RK4-adaptive superset is accepted as compliant. We report both compliance rates (Supplementary Table S1b) so that readers can judge directive adherence against the interpretation relevant to their setting.

##### S4.5 Recommended inspection script

The public dataset includes a helper (`evaluation/validate_run.py`) that, given a generated CSV, runs the following three checks and prints a pass/fail summary:

1. **File length check.** Is `csv_row_count` ≥ 7,200? If not, the run is Class D.
2. **Reference end-of-bolus check.** Is the bolus-peak concentration within 1 % of the reference value at t = 1 s? A failure flags bolus-handling bugs.
3. **Reference at-end check.** Is the concentration at t = 120 min within 1 % of the reference value? A failure flags unit mismatches and clearance bugs.

Applying this helper to our 200-run dataset catches all five defective runs within approximately 0.5 s per run.

#### Supplementary Appendix S5 — Recommended minimum validation protocol

For any user who plans to run an LLM-generated PK simulation script in a research or clinical context, we recommend the following minimum validation protocol before trusting the numerical output:

1. **Algorithm spot-check.** Read the source and confirm that the declared numerical method and step size match what was requested. Phase 2 of this study demonstrated that prompt-level directives are followed 98 % of the time (strict), but this is the baseline — not a guarantee.

2. **Unit audit.** Confirm explicitly that *time units are consistent throughout the script*. The three most damaging failures in this dataset were all caused by mixing minutes and seconds in a single integrator loop. Looking for `/ 60` or `* 60` conversions at the step-size level is the single highest-yield inspection.

3. **Event-timing audit.** Confirm that the loading dose is injected exactly once. The DeepSeek `p2_run14` duplicate-bolus bug would have been caught by a one-line check of the initial condition against the infusion loop's first iteration.

4. **Output-format sanity check.** Before trusting the output, read it: confirm that the CSV has the expected number of rows (7,201 for the 120-minute regimen at 1-second intervals) and that the first and last rows have sensible values.

5. **Numerical comparison against an independent reference.** Comparing three points — the bolus peak at t = 1 s, the end-of-infusion concentration at t = 60 min, and the end-of-simulation concentration at t = 120 min — against a validated reference (Tivatrainer, the reference simulator released with this paper, or equivalent) would have caught every defective run in this dataset. This step takes seconds and is the single most effective safeguard against silent numerical failure.

The public dataset includes a standalone validator (`evaluation/validation_triplet.py`) that implements exactly this check: pointed at any Marsh-model CSV produced under the standardized benchmark regimen, it reports per-point pass/fail against the reference values below and exits with a non-zero code if any point exceeds the 1 % tolerance.

##### S5.1 Minimum reference-based validation triplet: concrete values

For the standardized 70-kg / 140-mg-bolus / stepped-infusion regimen used throughout this benchmark, the three scalar comparisons described in step 5 above resolve to the following reference values:

| # | Check point | Reference $C_p$ ($\mu g \cdot mL^{-1}$) |
|---|---|---|
| 1 | Peak after bolus (t = 1 s) | 8.744 |
| 2 | End-of-infusion (t = 3,600 s) | 2.838 |
| 3 | Simulation horizon (t = 7,200 s) | 0.393 |

These values are taken from the reference simulator released with this paper (RK4 at dt = 0.01 s). Agreement within 1 % on all three points rules out every structural failure observed in the present dataset: unit mismatch (≥ 60-fold deviation at all three points), catastrophic Euler divergence (MDAPE ~ $10^5$ % throughout), duplicated bolus (approximately 100 % deviation at check point 1 — the bolus is effectively doubled, so $C_p$(t = 1 s) doubles), and header-only CSV (missing values at all three points).

#### S5.2 Reference validator (Python)

```python
# validation_triplet.py (simplified)
import pandas as pd

REFERENCE_POINTS = {
    1:    8.744,   # t = 1 s (peak after bolus)
    3600: 2.838,   # t = 3600 s (end of infusion)
    7200: 0.393,   # t = 7200 s (simulation horizon)
}
TOLERANCE_PCT = 1.0

def validate_triplet(csv_path):
    df = pd.read_csv(csv_path)
    for t_sec, ref_cp in REFERENCE_POINTS.items():
        row = df[df['time_sec'] == t_sec]
        if row.empty:
            return False  # header-only CSV or truncation
        pct_err = 100 * abs(row['Cp'].iloc[0] - ref_cp) / ref_cp
        if pct_err > TOLERANCE_PCT:
            return False
    return True
```

This validator flagged all 5 Class B/C/D runs in the present dataset and passed all 195 Class A runs, with observed errors on Class A runs bounded by 0.05 % at every check point.

#### S5.3 Scope

The triplet values above are specific to the Marsh model and to the regimen evaluated in this study. For Schnider, Eleveld, non-propofol models, or any regimen differing in bolus dose, patient weight, or infusion schedule, the reference values must be regenerated — either by running the open reference simulator under the desired configuration, or by extracting the same three time points from a validated clinical TCI tool such as Tivatrainer. The principle (three scalar comparisons at clinically meaningful time points against a validated reference within a tight tolerance) generalizes; the specific values do not.

### Supplementary Appendix S6 — Extended limitations and comparison with prior work

#### S6.1 Comparison with prior LLM-pharmacometrics studies

Shin et al. [1] evaluated ChatGPT and Gemini on NONMEM coding tasks and found that the models could generate initial templates but that these required expert revision due to structural and syntax errors. Our observation of discrete structural bugs in 2.5 % of runs is consistent with this: the failure modes are not random noise in the code, they are identifiable bugs that an expert reader would recognize and a naive user would not. Cloesmeijer et al. [2] report similar structural defects in R-based population PK workflows, including inconsistent unit handling — a direct parallel to our most frequent failure class.

Cha et al. [3] reported that ChatGPT succeeded on a three-compartment meropenem model with <1 % deviation from NONMEM but struggled with a covariate-dependent nivolumab model, with larger discrepancies attributed to incomplete covariate implementation. Our Marsh result of ~2.5 % failure rate on a fixed-parameter three-compartment model is therefore a conservative lower bound: when covariates are added (Schnider [8], Eleveld [9]), the additional implementation surface area exposes more opportunities for silent bugs.

Zheng et al. [6] examined seven OpenAI models (including o1 and GPT-4.1) on 13 NONMEM tasks and showed (a) degradation with task complexity in baseline conditions, and (b) substantial improvement with “optimized prompts”. Our Phase 2 result — that prompt-level specification of the integration method eliminates algorithmic error but does not eliminate unit or structural bugs — refines this: prompts can fix *algorithm selection* but cannot reach *numerical implementation details*.

Tosca et al. [12] provide a wider vision of how LLMs may contribute to pharmacometrics. Our work operationalizes the safety question embedded in that vision: what is the residual failure rate of the simplest task class, and what are the failure modes?

Shin and Ramanathan [13] show that prompt engineering can substantially raise the success rate of LLM-generated PK analysis code. Our single-shot 2.5 % failure rate should therefore be interpreted as an *upper bound* on the rate that would persist in a setting where users inspect generated code and iterate on errors. This iterative-refinement baseline is a natural target for future work.

Shahin et al. [14] propose multi-turn agentic workflows for quantitative clinical pharmacology, in which an LLM generates code, inspects its own output, and revises. Our benchmark provides the evaluation harness that such agentic workflows need: quantitative accuracy metrics tied to clinical validation criteria.

#### S6.2 Study-design limitations

**Single-shot generation.** We deliberately used a single-shot protocol with no iterative error correction. In practice, many users inspect the code, test it, and paste errors back to the LLM. Our observed 2.5 % failure rate should therefore be read as an upper bound on the rate that would persist after one round of human review [13].

**Default, non-reasoning response mode.** We used each vendor's default chat mode, reflecting the most common end-user interaction pattern. Dedicated reasoning/thinking variants (OpenAI o-series, Claude's extended thinking, Gemini's deep-thinking) are likely to perform better and are a natural target for follow-up work.

**Five LLMs only.** The sample is small. Every Class C numerical failure and the Class B duplicated-bolus run came from DeepSeek or Grok, while the single Class D execution failure came from Gemini; this may reflect training-data coverage, post-training on scientific-computing tasks, or base-model scale differences, but our study cannot distinguish among these. This observation is hypothesis-generating rather than inferential.

**Marsh only.** As stated in the main text, Marsh is the simplest fixed-parameter three-compartment propofol model. Extending this benchmark to Schnider and Eleveld is planned future work; the open dataset release is designed to support such extensions directly.

**Prompt engineering not systematically explored.** Our Phase 1 / Phase 2 comparison is a two-point intervention study. Systematic prompt engineering — instruction chaining, few-shot examples, self-consistency prompting — is not studied here. Shin and Ramanathan [13] provide initial evidence that prompt engineering can raise success rates substantially.

**No iterative error-feedback loops.** Many production workflows for LLM code generation involve a human or an automated agent inspecting the output and feeding errors back to the LLM [14]. This iterative mode is known to improve code quality substantially and is out of scope here.

**Rank-based statistics.** The Kruskal–Wallis, Dunn, and Mann–Whitney tests we use cannot distinguish "catastrophically wrong" from "mildly wrong" because they operate on ranks. This is why we supplement the rank-based analyses with Varvel-style ordinal classification.

#### Supplementary Appendix S7 — AI use disclosure

The use of AI tools during manuscript preparation is disclosed below.

**AI tool used.** Claude Sonnet 4.6 (Anthropic), accessed through the Claude.ai web interface and Claude Code command-line interface (October 2025 through April 2026).

**Tasks for which AI assistance was used:** 1. Code review of the reference simulator (`reference/reference.py`) for numerical-methods correctness. 2. Construction of the analysis pipeline (`evaluation/`), including the per-run PE computation, the Kruskal–Wallis/Dunn/Mann–Whitney pipeline using `scipy.stats` and `scikit-posthocs`, and the figure-generation scripts. 3. Drafting assistance for the manuscript text, including the structural reorganization of sections, rewording for concision, and consistency checks between the English and Japanese versions. 4. Structural review of the final manuscript PDFs for preprint deposition and journal submission.

**Tasks performed without AI assistance:** 1. Design of the benchmark protocol, including Phase 1 / Phase 2 definition, the choice of Marsh parameters, the dosing regimen, and the selection of MDPE/MDAPE/Wobble as primary metrics. 2. All LLM code-generation runs and their execution (the LLMs being evaluated were treated as the subject of the study, not as tools). 3. Statistical analysis design and interpretation. 4. All scientific conclusions.

**Verification.** Every AI-assisted output was independently verified by the author before inclusion: generated code was executed and compared against the reference, generated text was read and edited, and all numerical results were recomputed from raw data by the author. The author takes full responsibility for all content.

#### Supplementary Table S1. LLM model versions, access dates, and account configurations

| LLM | Exact model string / version | Interface | Access period | Account tier | Reasoning mode |
|---|---|---|---|---|---|
| ChatGPT | GPT-5.3 Instant | chatgpt.com web interface | 2026-02 through 2026-04 | Plus (paid) | auto-route disabled; non-reasoning |
| Claude | Claude | claude.ai web | 2026-02 | Pro (paid) | non- |

| LLM | Exact model string / version | Interface | Access period | Account tier | Reasoning mode |
|---|---|---|---|---|---|
| | Sonnet 4.6 (claude-sonnet-4-6-20260105) | interface | through 2026-04 | | reasoning (standard mode) |
| DeepSeek | DeepSeek-V3.2 | chat.deepseek.com web interface | 2026-02 through 2026-04 | Free tier | Instant mode (non-reasoning) |
| Gemini | Gemini 3 (displayed as "Fast" in the web UI) | gemini.google.com web interface | 2026-02 through 2026-04 | Google AI Pro (paid) | non-reasoning (Fast variant) |
| Grok | Grok 4.1 Fast | grok.com web interface | 2026-02 through 2026-04 | SuperGrok (paid) | Fast mode (non-reasoning) |

All five LLMs were accessed through vendor-provided public chat interfaces (no API access). The default non-reasoning response mode was selected in every case to reflect the most common end-user interaction pattern. Reasoning/Thinking variants (OpenAI o-series, Claude Extended Thinking, DeepSeek-R1, Gemini Pro with Deep Thinking, Grok Heavy) were deliberately not used; their evaluation is planned future work.

#### Supplementary Table S1b. Phase 2 compliance with prompt directives

| LLM | n | Algorithm declared | Step size declared | Strict RK4 compliance | Lenient RK4 compliance | dt = 1 s compliance |
|---|---|---|---|---|---|---|
| ChatGPT | 20 | Classical RK4: 20 | 1.0 s: 20 | 100 % (20/20) | 100 % (20/20) | 100 % (20/20) |
| Claude | 20 | Classical RK4: 20 | 1.0 s: 20 | 100 % (20/20) | 100 % (20/20) | 100 % (20/20) |
| DeepSeek | 20 | Classical RK4: 18, RK4-adaptive: 2 | 1.0 s: 20 | 90 % (18/20) | 100 % (20/20) | 100 % (20/20) |
| Gemini | 20 | Classical RK4: 20 | 1.0 s: 20 | 100 % (20/20) | 100 % (20/20) | 100 % (20/20) |

| LLM | n | Algorithm declared | Step size declared | Strict RK4 compliance | Lenient RK4 compliance | dt = 1 s compliance |
|---|---|---|---|---|---|---|
| Grok | 20 | Classical RK4: 20 | 1.0 s: 20 | 100 % (20/20) | 100 % (20/20) | 100 % (20/20) |
| **Total** | **100** | Classical RK4: 98, RK4-adaptive: 2 | 1.0 s: 100 | **98 % (98/100)** | **100 % (100/100)** | **100 % (100/100)** |

Strict compliance counts only classical RK4 as compliant. Lenient compliance additionally accepts RK4 with adaptive step-size correction (a strict superset of classical RK4) as compliant with the prompt's directive for fourth-order Runge–Kutta. The DeepSeek Phase 2 runs 17 and 19 implemented RK4-adaptive and produced numerically excellent output (both Class A, MDAPE < 0.01 %); they are non-compliant under the strict interpretation and compliant under the lenient interpretation. See Supplementary Appendix S4.4 for details.

#### Supplementary References

Supplementary reference numbering is self-contained within this appendix and mirrors the main text for cross-referenced items.


1. Shin E, Yu Y, Bies RR, Ramanathan M. Evaluation of ChatGPT and Gemini large language models for pharmacometrics with NONMEM. J Pharmacokinet Pharmacodyn 2024;51:187–197. DOI: 10.1007/s10928-024-09907-w.

2. Cloesmeijer ME, Janssen A, Koopman SF, Cnossen MH, Mathôt RAA; SYMPHONY consortium. ChatGPT in pharmacometrics? Potential opportunities and limitations. Br J Clin Pharmacol 2024;90:360–365. DOI: 10.1111/bcp.15895.

3. Cha HJ, Choe K, Shin E, et al. Leveraging large language models in pharmacometrics: evaluation of NONMEM output interpretation and simulation capabilities. J Pharmacokinet Pharmacodyn 2025;52:34. DOI: 10.1007/s10928-025-09956-9.

4. Zheng W, Wang W, Kirkpatrick CMJ, Landersdorfer CB, Yao H, Zhou J. AI for NONMEM coding in pharmacometrics research and education: shortcut or pitfall? CPT Pharmacometrics Syst Pharmacol 2025;14:1965–1969. DOI: 10.1002/psp4.70125.

5. Marsh B, White M, Morton N, Kenny GNC. Pharmacokinetic model driven infusion of propofol in children. Br J Anaesth 1991;67:41–48. DOI: 10.1093/bja/67.1.41.

6. Schnider TW, Minto CF, Gambus PL, et al. The influence of method of administration and covariates on the pharmacokinetics of propofol in adult volunteers. Anesthesiology 1998;88:1170–1182. DOI: 10.1097/00000542-199805000-00006.

7. Eleveld DJ, Colin P, Absalom AR, Struys MMRF. Pharmacokinetic–pharmacodynamic model for propofol for broad application in anesthesia and sedation. Br J Anaesth 2018;120:942–959. DOI: 10.1016/j.bja.2018.01.018.

8. Tosca EM, Aiello L, De Carlo A, Magni P. Pharmacometrics in the age of large language models: a vision of the future. Pharmaceutics 2025;17:1274. DOI: 10.3390/pharmaceutics17101274.

9. Shin E, Ramanathan M. Evaluation of prompt engineering strategies for pharmacokinetic data analysis with the ChatGPT large language model. J Pharmacokinet Pharmacodyn 2024;51:101–108. DOI: 10.1007/s10928-023-09891-7.

10. Shahin MH, Goswami S, Lobentanzer S, Corrigan BW. Agents for change: artificial intelligent workflows for quantitative clinical pharmacology and translational sciences. Clin Transl Sci 2025;18:e70188. DOI: 10.1111/cts.70188.
